# Deep learning-based prognosis models accurately predict the time to delivery among preeclampsia patients using health records at the time of diagnosis

**DOI:** 10.1101/2022.04.03.22273366

**Authors:** Xiaotong Yang, Hailey K Ballard, Aditya D Mahadevan, Ke Xu, David G Garmire, Elizabeth S Langen, Dominick J Lemas, Lana X Garmire

**Author notes:** Corresponding author: Lana Garmire Room 3366, Building 520, NCRC, 1600 Huron Parkway, Ann Arbor, MI 48105.

## Abstract

**Background:** Preeclampsia (PE) is one of the leading factors in maternal and perinatal mortality and morbidity worldwide with no known cure. Delivery timing is key to balancing maternal and fetal risk in pregnancies complicated by PE. Delivery timing of PE patients is traditionally determined by closely monitoring over a prolonged time. We developed and externally validated a deep learning models that can predict the time to delivery of PE patients, based on electronic health records (EHR) data by the time of the initial diagnosis, in the hope of reducing the need for close monitoring.

**Method:** Using the deep-learning survival model (Cox-nnet), we constructed time-to-delivery prediction models for all PE patients and early-onset preeclampsia (EOPE) patients. The discovery cohort consisted of 1,533 PE cases, including 374 EOPE, that were delivered at the University of Michigan Health System (UM) between 2015 and 2021. The validation cohort contained 2,172 PE cases (547 EOPE) from the University of Florida Health System (UF) in the same time period. We built clinically informative baseline models from 45 pre-diagnosis clinical variables that include demographics, medical history, comorbidity, PE severity, and initial diagnosis gestational age features. We also built full models from 60 clinical variables that include additional 15 lab tests and vital signs features around the time of diagnosis.

**Results:** The 7-feature baseline models on all PE patients reached C-indices of 0.74 and 0.73 on UM hold-out testing and UF validation dataset respectively, whereas the 12-feature full model had improved C-indices of 0.79 and 0.74 on the same datasets. For the more urgent EOPE cases, the 6-feature baseline model achieved C-indices of 0.68 and 0.63, and its 13-feature full model counterpart reached C-indices of 0.76 and 0.67 in the same datasets.

**Conclusions:** We successfully developed and externally validated an accurate deep-learning model for time-to-delivery prediction among PE patients at the time of diagnosis, which helps to prepare clinicians and patients for expected deliveries.

## Introduction

Preeclampsia (PE) is a pregnancy complication affecting 2% to 8% of all pregnancies worldwide and is a leading cause of maternal, fetal, and neonatal mortality and morbidity^1,2^. PE is defined by new-onset hypertension after 20 weeks of gestation and the presence of proteinuria, and/or other signs of end-organ damage. PE is a diverse syndrome with various subtypes along the spectrum of gestational hypertensive disorders^3^. It can be divided into early-onset PE (diagnosed before 34 weeks of pregnancy) or late-onset PE (diagnosed after 34+0 weeks of pregnancy); PE with severe features (sPE) or PE without severe features^4,5^. Failure to properly manage PE can lead to a wide variety of severe maternal and neonatal adverse outcomes according to the iHOPE study, while the only known cure for PE is delivery of the placenta^6–8^. Although earlier delivery can significantly reduce the risk of maternal adverse outcomes, it is associated with increased neonatal unit admission among preterm patients. This, especially in cases of EOPE ^9^, creates a dilemma as earlier delivery can potentially prevent severe morbidities including maternal seizure, stroke, organ dysfunction, and intrauterine fetal demise, but may lead to premature birth and subsequent neonatal complications^10,11^. To balance the risks to both mother and baby, current clinical management of PE includes supportive blood pressure management and prophylaxis for maternal seizures, and a two-dose intramuscular course of betamethasone to augment fetal lung maturation^12^.

Generally, delivery is recommended for PE patients with more than 37+0 weeks of gestation and for severe PE patients with more than 34+0 weeks of gestation^12^. In reality, the delivery timing is a more complex problem, clinicians need to consider both the fetal development, maternal and fetal risk of complications, and availability of ICU resources when deciding on delivery timing, particularly among challenging EOPE cases^12,13^. The decision of delivery is usually made after close monitoring and extensive testing on preeclampsia patients over a prolonged time, which may not be easily accessible and affordable to all patients (particularly those in rural areas or under-developed countries). In addition, current risk assessment tools focus on maternal risk prediction but not the overall delivery urgency considering both moms and fetuses. FullPIERS, miniPIERS and PREP-S are well-established and externally validated models to predict the maternal risk of adverse outcomes among PE patients, in the hope of assisting delivery decisions^14–19^. These tools are recommended by some, but not all national guidelines^12,13^. Most of these tools only predict maternal risks, however, clinicians need to consider both maternal and neonatal outcomes when deciding when to deliver. A patient at 34 weeks of gestation would have very different delivery timing compared to a patient at 37 weeks of gestation, even if they have the same risk of adverse outcomes. It is of great importance to directly and precisely predict the time to delivery as early as the first diagnosis of PE, which allows the clinicians to assess the delivery urgency early on and to help them better prioritize resources and treatment, particularly for those doctors practicing in rural or under-developed countries. Additionally, the aforementioned risk predictor models do not assess the risk from baseline features, such as the patient’s race, social status, lifestyle, and other comorbidities, which may also have influences on delivery timing.

Toward this goal, we developed and externally validated the first deep learning model to predict patient delivery time after the initial diagnosis of PE using electronic health records (EHR) data. We utilized the state-of-the-art deep learning-based prognosis prediction model, Cox-nnet (version 2), which we previously developed ^20–22^. Cox-nnet methods had previously consistently shown excellent predictive performances under a variety of conditions, including on EHR data^20^. Our objectives were: (1) to predict the time to delivery at the first diagnosis of PE for all PE patients and an EOPE sub-cohort, by constructing and validating deep-learning models utilizing EHR data; and (2) to assess the quantitative contributions of critical EHR features informative of delivery time among PE patients, including those EOPE patients.

## Methods

### Data Source

We obtained the discovery cohort of the Precision Health Initiative of Michigan Medicine (UM), the academic healthcare system of the University of Michigan^23^. Data usage was approved by the Institutional Review Board (IRB) of the University of Michigan Medical School (HUM#00168171). We obtained the validation cohort from the Integrated Data Repository database at the University of Florida (UF). Data usage was approved by IRB of the University of Florida (#IRB201601899). In both cohorts, we extracted all obstetric records with at least one PE diagnosis between 2015 to 2021 based on ICD-10 diagnosis codes (**Supplementary Table 1**). We excluded patients with the following conditions: Hemolysis, Elevated Liver Enzymes, and Low Platelet (HELLP) syndrome and eclampsia, for which iatrogenic delivery is ubiquitously induced within 48 hours of diagnosis despite fetal condition; chronic hypertension with superimposed PE, whose onset may occur before week 20 and with no clear definitions in the United States^24^; and postpartum PE, which is only developed after delivery. We also removed patients transferred from other institutions by deleting patients with no visit record within 180 days before the first diagnosis of PE to ensure the accuracy of the initial diagnosis time of PE. The resulting discovery cohort consisted of 1,533 PE cases (including 374 EOPE cases) and the validation cohort contained 2,172 PE cases (including 547 EOPE).

### Fully connected Cox-nnet neural network models

We constructed all models using the Cox-nnet v2 algorithm (**Supplementary Figure 1**)^20^. In this study, we adopted the model to predict the time between PE diagnosis to delivery. To ensure the stability of the models, we divided the discovery dataset into a training set (80%) and a hold-out testing set (20%) and applied 5-fold cross-validation on the training set.

### EHR Feature Engineering

We extracted all available features from UM Precision Health Initiative EMR data. We developed 4 models to predict the time to delivery of PE patients: PE baseline, PE full, EOPE baseline and EOPE full models. As suggested by clinicians, the initial baseline models include demographics, medical history, comorbidities, the severity of PE, pregnancy and fetal development characteristics. The full model incorporated all features from the baseline model, with additional lab results and vital signs commonly collected within 5 days before the initial diagnosis of PE (**Supplementary Figure 2A**). EOPE models were built and tested using the same features on patients with PE onset time before 34 weeks of gestation. Features with low powers and high correlation were removed to ensure model accuracy.

Pregnancy characteristics included parity, number of fetuses, gestational age, PE severity at initial diagnosis, and history of preterm birth, c-section, abruption, etc. Fetal development includes poor fetal growth according to the associated ICD code(O36.59). Other comorbidities were grouped into 29 categories using the Elixhauser Comorbidity Index^1^. The observational window for lab results and vital signs was 5 days before the day of the initial PE diagnosis. Only the first results of repeated lab tests were used to avoid intervention/treatment effects. Summary statistics of systolic blood pressure (SBP), diastolic blood pressure (DBP), and respiratory rate (RR) measures were included (max, min, mean, standard deviation), as done in previous work^2^. We removed features with high missing proportions (over 20%) and sparse features with fewer than 10 non-zero values. Highly correlated variables were identified using the variance inflation factor (VIF) and removed one at a time until all features had a VIF below 3 to avoid multicollinearity. The remaining missing values were imputed using the PMM algorithm from R package “mice”. All numerical features were scaled by dividing their root mean square. Numeric features with skewness above 3 were log-transformed. As a result, 60 features were kept for initial analysis (**Supplementary Table 2, Supplementary Figure 2B).**

### Reduced feature representation from the Cox-nnet models

To derive a subset of clinically significant and easily interpretable features, we reduced Cox-nnet features based on both their importance scores and significance levels. To do so, we first selected the top 15 (25% of total features) most important features based on their average permutation importance scores generated by Cox-nnet models. Permutation important scores provide more stable results than other feature selection methods on this dataset, including stepwise selection, lasso regularization, and random forest feature selection^25^. Then we calculated the log-rank p-value for the 15 features individually and selected the significant ones. We also conducted the ANOVA test on the remaining features to ensure their powers (**Supplementary Table 3**). We rebuilt the clinically informative Cox-nnet models with the reduced set of features, the same way as the models using all initial input features.

### Model evaluation

We evaluated the cross-validation, hold-out test, and validation results of each model using Harrel’s concordance index (C-index). The C-index evaluates the accuracy of predicted events by comparing their relative order to the order of actual events. It is frequently used to assess survival predictions^26^. The reported C-indices in the training data are the repeated results of the 5-fold cross-validation C-indices on the training sets. To enhance the interpretation of the prognosis prediction, we also stratified patients into high, medium, and low-urgency groups based on the predicted results plotted the Kalper-Meier (KM) curves of time-to-delivery in each group and reported the log-rank p-values. The log-rank test, on the other hand, compares the survival distribution between patient groups, assuming no differences in survival exist^27^(p4). Additionally, we used each clinically informative and reduced model result to predict the chances of patients delivering within 2 days, 7 days, and 14 days and obtained the AUROC (area under the receiver operating curve) for each task.

### Interactive Web Application for Easy Model Validation

To disseminate the models for public use, we containerized the pre-trained Cox-nnet model into a Docker-based web application using R shiny^28^. This allows the users to access the models easily through a local web interface and get prediction results quickly. This app contains two main panels: the individual prediction panel and the group prediction panel. Using pre-trained models, the individual prediction panel calculates the prognosis index (PI) score of a single new patient, marking its positions and percentiles in a distribution plot of PIs within the UM discovery cohort. The group panel takes in a group of new patients and returns predicted PIs and percentiles of their PIs in a table. The shiny app is available at http://garmiregroup.org/PE-prognosis-predictor/app

### External Validation using UF data

We validated the reduced models on a large external EHR dataset from the University of Florida. We extracted and processed the same features included in the baseline, full, EOPE-baseline and EOPE-full model (see Methods). The authors uploaded cleaned UF data to the shiny app described above, and the app automatically produced predicted values using the packaged models trained on UM dataset. The development and validation strictly followed the TRIPOD checklist(**Supplementary Table 4**).

### Estimate time-to-deliver using maternal risk of adverse outcomes calculated from the fullPIERS model

The fullPIERS model is a model to predict the maternal risk of adverse outcomes in PE patients, yet it cannot effectively predict time-to-delivery at the initial diagnosis of PE. To illustrate this, we calculated the maternal risk of adverse outcomes using the fullPIERS formula as reported by von Dadelszen et al^14^, used this risk score to estimate the time-to-delivery and compared its performance with our proposed model.

We estimated the probability of adverse outcomes (p) and calculated its concordance index with time to deliver for all PE patients and EOPE subsets, following the original paper. We also plotted the survival curves of high-risk (top 25%), middle-risk (25% - 75%) and low-risk (bottom 25%) groups for all PE and EOPE patients. One limitation is that we do not have chest pain/dyspnoea or SpO2 information collected, so we assume no patients have chest pain or dyspnoea and all patients have 97% SpO2, as instructed by the FullPIERs web calculator https://pre-empt.obgyn.ubc.ca/home-page/past-projects/fullpiers/.

### Software

R 4.2.1 and Python were used for all analyses^29,30^. R package “dplyr”, “mice” were used in data preparation^31,32^. R package “shiny” and continuumio/anaconda3 Docker image were used to build an interactive web application^28^. Python version 3.9 and R version 4.2.1 are used to run the models in the Docker containers.

## RESULT

### Cohort characteristics

The discovery cohort consisted of 1,533 PE cases, including 374 EOPE cases collected from the University of Michigan Precision Health and the validation cohort contained 2,172 PE cases (including 547 EOPE) collected from the University of Florida Health System between 2015 and 2022. (**Figure 1**). We employed their EHR data to predict their time-to-delivery from the initial diagnosis of PE. Summaries of the patient characteristics of these cohorts are shown in **Table 1** and **2**.

**Figure 1:**
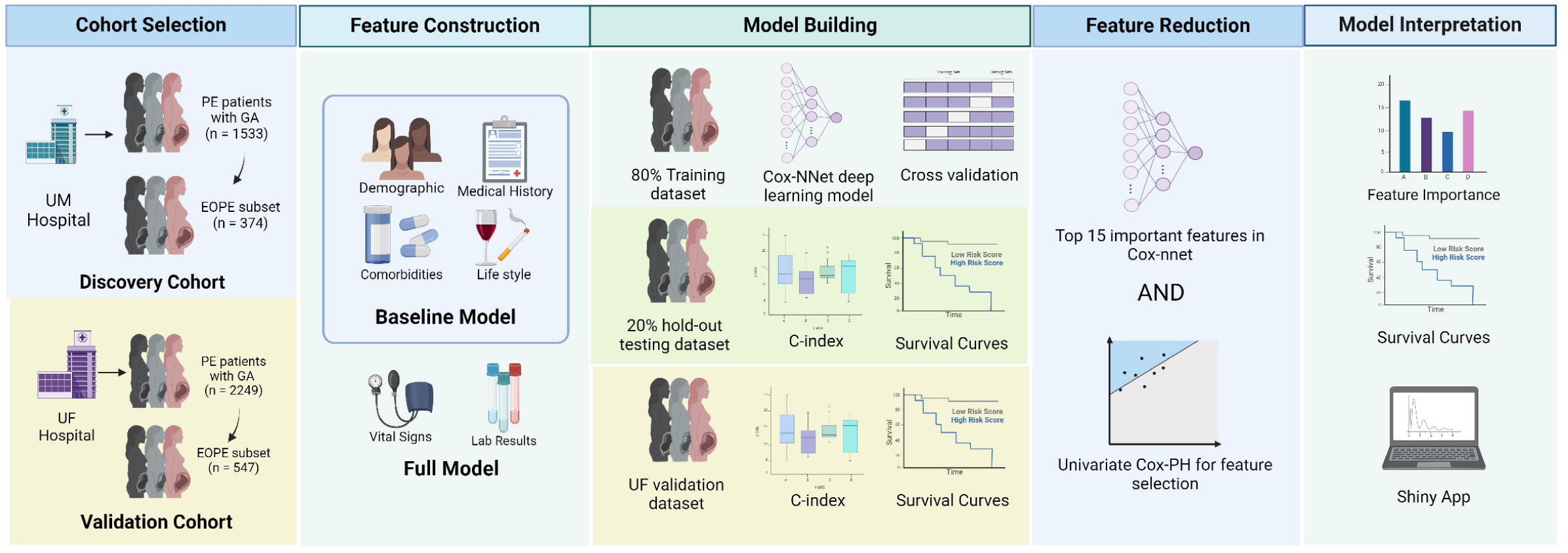
Experimental design and cohort overview. **A) Experiment Design Workflow**: The discovery cohort was obtained from the University of Michigan Health System and a validation cohort of similar size and time was obtained from the University of Florida Health System. We constructed 4 predictive models: baseline and full models for all PE patients and baseline and full models for EOPE patients. The input variables in baseline models include patients’ demographics, lifestyle, comorbidities and medical history. The full models include additional lab tests and vital signs from within 5 days of PE diagnosis, in addition to the variables in the baseline models. We trained the Cox-nnet prognosis prediction model using 80% training from the discovery cohort, tested it on 20% hold-out data from the discovery cohort, and validated it using the validation cohort. We then built clinically informative models by reducing Cox-nnet features based on both their importance scores and significance levels. The models are examined by the importance scores of top features and stratified survival curves based on patient survival risks. We disseminated the feature-reduced, clinically informative models into a user-friendly web application for healthcare professionals to use. Created with <u>BioRender.com</u>.

**Table 1:** Basic Patient Characteristics for this study.

| *Variable Name* | *University of Michigan PE*<br>*Discovery Cohort (n = 1533)* | *University of Florida PE*<br>*Validation Cohort (n = 2172)* |
| --- | --- | --- |
| ***Maternal Age, mean (SD)*** | 30.19(5.74) | 28.63(6.46) |
| ***Parity, mean (SD)*** | 0.72(1.15) | 0.73(1.55) |
| ***Number of fetuses, mean (SD)*** | 1.07(0.27) | 1.04(0.21) |
| ***Gestational Age at Diagnosis***<br>***(days), mean (SD)*** | 249.96(27.44) | 256.72(26.83) |
| ***Time to Delivery (days), mean***<br>***(SD)*** | 7.02(15.39) | 5.70(13.28) |
| ***Race, N (%)*** |  |  |
| *African American* | 0.18 | 0.37 |
| *American Indian or Alaska Native* | 0.00 | 0.00 |
| *Asian* | 0.07 | 0.02 |
| *Caucasian* | 0.75 | 0.49 |
| *Native Hawaiian and Other Pacific*<br>*Islander* | 0.00 | 0.00 |
| *Unknown or Other* | 0.00 | 0.12 |
| ***Ethnicity, N (%)*** |  |  |
| *Hispanic* | 0.06 | 0.11 |
| *Non-Hispanic* | 0.94 | 0.87 |
| *Unknown* | 0.00 | 0.01 |
| ***Smoking Status, N (%)*** |  |  |
| *Current Smoker* | 0.06 | 0.13 |
| *Former Smoker* | 0.24 | NA |
| *Never Smoker* | 0.70 | 0.87 |
| ***Illegal Drug Use Status, N(%)*** |  |  |
| *Yes* | 0.10 | 0.17 |
| *No* | 0.90 | 0.83 |
| ***History of PE, N (%)*** |  |  |
| *Yes* | 0.11 | 0.12 |
| *No* | 0.89 | 0.88 |
| ***sPE, N (%)*** |  |  |
| *Yes* | 0.47 | 0.35 |
| *No* | 0.53 | 0.65 |

### The baseline prediction model of time to delivery interval among PE patients

PE is a syndrome with well-characterized phenotypes, where hypertension is the most significant clinical symptom. Thus the structured data in the EHR system provide the most useful and straightforward information. From the structured data, we obtained 45 variables including patient demographics, medical history, comorbidities, PE diagnosis time, and severity after data preprocessing (**Supplementary Table 2**). The resulting model has very decent performance with C-indices of 0.73, 0.72, and 0.71 in the UM cross-validation, UM hold-out testing, and UF validation cohorts, respectively (**Figure 2A**).

**Figure 2:**
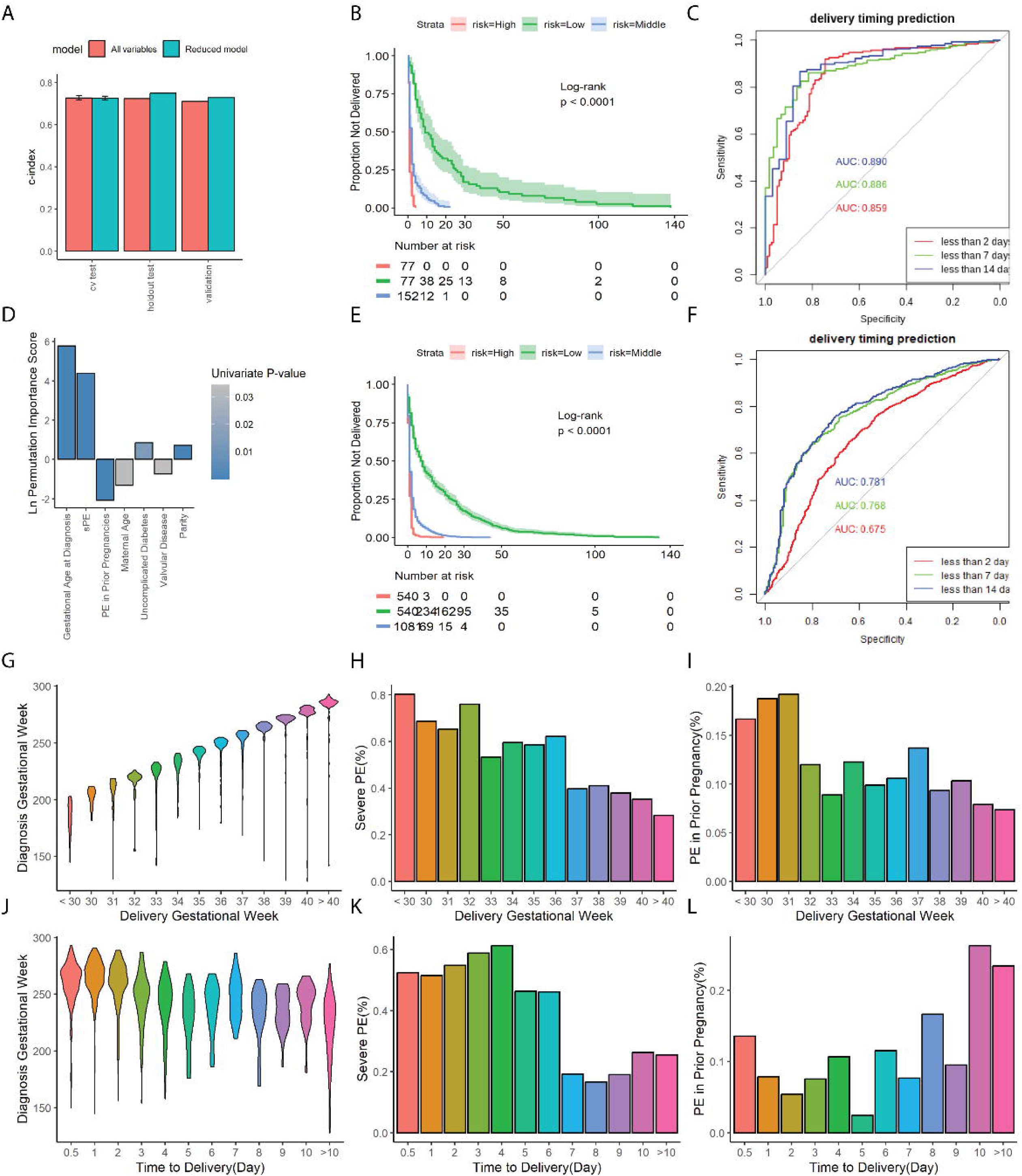
PE Baseline model results, interpretation, and evaluation. A: The bar plots of C-indices from the original Cox-nnet models (red) and feature-reduced clinically informative model (green), on the UM cross-validation and hold-out testing set and UF validation set. B, E: The survival curves of high-risk (top 25%), intermediate-risk (middle 50%) and low-risk groups (bottom 25%), categorized by predicted PI from the reduced baseline model in A on B: hold-out testing data and E: validation data. C, F: ROC curves of prediction delivery time within 2 days, 7 days and 14 days using results from reduced baseline model on C: hold-out testing data and F: validation data. D: The ln-transformed permutation importance scores of features in the feature-reduced baseline model. A positive sign indicates that a higher value in feature is associated with a shorter time to delivery and a negative sign means an extension of time to delivery. G-L: The distribution of diagnosis gestational age, sPE rate and PE in prior pregnancy rate, in associations with delivery gestational week (G-I) and time (days) to delivery (J-L).

To enhance the clinical utilities of the Cox-nnet model, we reduced the number of predictive features following the feature reduction procedure in the Method section. This procedure resulted in 7 significant features, which we used to rebuild the “clinically informative Cox-nnet baseline model”. It has C-index scores of 0.73, 0.74, and 0.73 on UM cross-validation, hold-out testing, and UF validation dataset respectively (**Figure 2A**). We stratified patients into 3 groups by the quartiles of predicted time-to-delivery from the reduced model: high-risk (upper quartile), intermediate-risk (interquartile), and low-risk (lower quartile) groups. The survival curves of the time to delivery interval on these three risk groups display significant differences (log-rank p-value < 0.0001) on both the hold-out testing set (**Figure 2B**) and validation set (**Figure 2E**), confirming the strong discriminatory power of the PI score. To enhance the interpretability of the prognosis modeling, we stratified this model using the threshold of 2/7/14 days and predicted the accuracies of delivery using these classifications. The AUROC scores of these classification tasks are 0.85, 0.88, and 0.89 on the testing set **(Figure 2C)** and 0.67, 0.76, and 0.75 on the validation set **(Figure 2F**), respectively.

The seven features in the clinically informative baseline model included those that shorten the time to delivery and extend the time to delivery (**Figure 2D**; **Table 3**). In descending order of importance scores, the features that shorten the time to delivery are gestational age at diagnosis, sPE, uncomplicated pregestational diabetes mellitus, and parity. Conversely, features extending the time to delivery are PE in a prior pregnancy, increasing maternal age, and comorbid valvular disease. To demonstrate the associations of these important features with time to delivery, we dichotomized patient survival in the hold-out testing set by the median value of each feature (**Supplementary Figure 3**). All features, except maternal age, show significant differences (log-rank p-value < 0.05) between the dichotomized survival groups. We further examined the relationship of the top 3 features (gestational age at diagnosis, sPE, and history of PE in prior pregnancy) with the gestational age at delivery and time to delivery (day) using the UM discovery set in (**Figure 2G-2L).** Later gestational age at diagnosis leads to a later gestational age of delivery (**Figure 2G**), but a shorter time to delivery (**Figure 2J**). sPE is associated with earlier gestational age of delivery (**Figure 2H**) and shorter time to delivery (**Figure 2K**) are diagnosed with sPE. In the deliveries from smaller (<32 weeks) gestational ages, the percentages of patients with PE in prior pregnancies are significantly higher (**Figure 2I)**. However, the percentages of prior PE fluctuate with respect to time to delivery (**Figure 2L**).

Worth noticing, that not all patients diagnosed with PE in 37 weeks or later delivered the babies right away, despite being the least severe cases and can be delivered quickly according to the medical recommendation^12^. Nevertheless, we alternatively built another baseline model with only those patients diagnosed before 37 weeks of gestation. We observed very similar results as the above baseline model using all PE patients, in terms of C-index, the selected top features and their feature scores (**Supplementary Figure 4 A-D**).

### The full model of time to delivery among PE patients

We next investigated the contribution to time of delivery from all 60 variables, including the 45 baseline variables above and an additional 15 laboratory testing results and vital signs obtained in the 5-day observation window before the time of diagnosis (**Supplementary Table 2**). The clinical informative model after feature reduction consists of 12 top features (**Table 3**). This model shows significantly (P< 0.001, t-test) higher cross-validation accuracy of time to delivery compared to the seven-feature baseline model, with median C-index scores almost as high as 0.80, with 0.78, 0.79, and 0.74 in the cross-validation, testing, and validation datasets respectively. These C-indices are excellent for survival predictions, despite the high heterogeneity of PE and the large patient size which makes it difficult to predict delivery time precisely^18,33^. The Kaplan-Meier curves of the high-, intermediate- and low-risk groups show more significant distinction in testing (**Figure 3B)** and validation set (**Figure 3E**), than the baseline model (**Figure 2B and 2E**). Similarly, we stratified the full model using the threshold of 2/7/14 days and predicted the accuracies of delivery using these classifications. The AUROC scores of these classification tasks are 0.88, 0.93, and 0.93 on the testing set and 0.84, 0.89, and 0.90 on the validation set respectively **(Figure 2C, 2F**).

**Figure 3:**
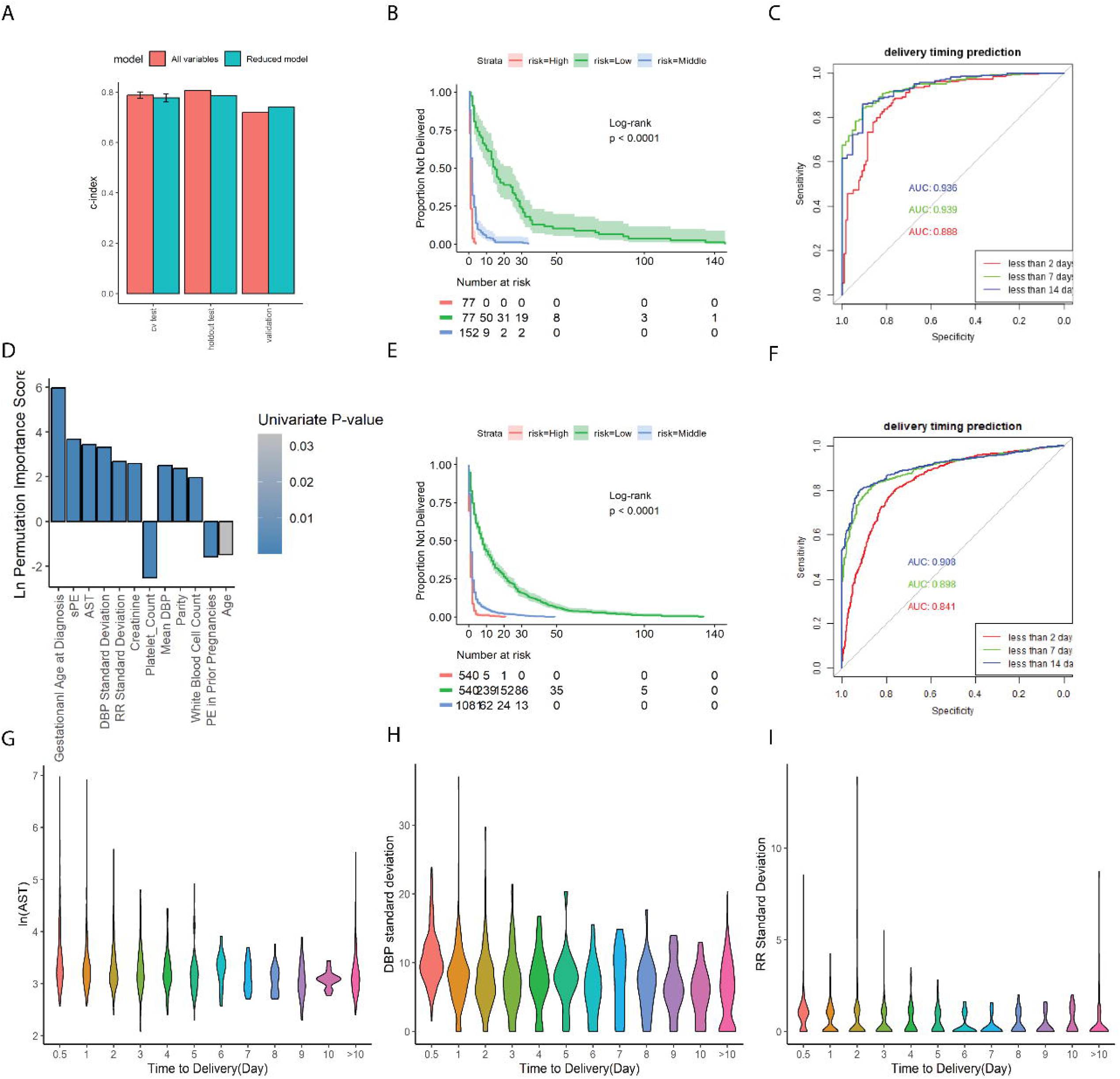
PE Full model results, interpretation and evaluation. A: The bar plots of C-indices from the original models (red) and feature-reduced clinical informative model (green), on the UM training cross-validation and hold-out testing set and UF validation set. B, E: The survival curves of high-risk (top 25%), intermediate-risk (middle 50%) and low-risk groups (bottom 25%), categorized by predicted PI from the reduced full model in A. B: hold-out testing data, E: validation data. C, F: ROC curves of prediction delivery time within 2 days, 7 days and 14 days using results from reduced full model (A) on C: hold-out testing data and F: validation data. D: The ln-transformed permutation importance scores of features in the feature-reduced baseline model. A positive sign indicates that a higher value in the feature is associated with a shorter time to delivery and a negative sign means an extension of time to delivery. G-I: The distribution of aspartate aminotransferase (AST) values, the standard deviation of diastolic blood pressure (DBP) and the standard deviation of respiratory rate (RR), in association with time (days) to delivery.

Further examination of the 12 important features in the full model (**Figure 3D**, **Table 3**) shows good consistency with the 7-feature baseline model (**Figure 2D**, **Table 3)**. Five out of seven features in the baseline model also exist in the full model with similar importance scores: gestational age at diagnosis, sPE, parity, maternal age, and PE in prior pregnancies. Gestational age at PE diagnosis and sPE continued to be the two most important features in the full model. We also identify new important features from lab tests and vital signs: aspartate aminotransferase (AST) value, the standard deviation of diastolic blood pressure (DBP), the standard deviation of respiratory rate (RR), creatinine value, mean DBP and white blood cell count (**Figure 3D**). Conversely, platelet count is a new feature with a negative importance score, associated with a longer time to delivery. All dichotomized survival plots using median stratification on each of the 12 important features have log-rank p-values smaller than 0.05, confirming their associations with time to delivery in the discovery set (**Supplementary Figure 5)**. We examined the 3 top lab/vital sign features: AST, the standard deviation of DBP, and the standard deviation of RR, on their association with the duration of time between diagnosis and delivery. These values show negative trends with time to delivery, particularly for AST value and the standard deviation of DBP (**Figure 3G-I**). These 3 features are roughly uniformly distributed across delivery gestational ages, except AST which shows slightly higher values in deliveries less than 32 weeks of gestational age (**Supplementary Figure 6)**.

Similar to the baseline model earlier using PE patients diagnosed before 37 weeks of gestation, we again alternatively built another full model with the same patients before 37 weeks of gestation. We observed very similar results as the full model using all PE patients, in terms of C-index, the selected top features and their feature scores (**Supplementary Figure 4 E-H**).

### Time to delivery prediction of EOPE patients

Accurate prediction of EOPE patients’ time to delivery is crucial, given that delivery of a premature infant has more significant neonatal consequences. Using similar modeling techniques, we trained two additional EOPE-specific Cox-nnet v2 models (baseline vs. full model), using the same features described earlier (**Supplementary Table 2**), on a subset of 374 EOPE patients from the UM discovery cohort.

The C-indices for the clinically informative EOPE baseline model are 0.67, 0.68, and 0.63 on the UM cross-validation, hold-out testing, and UF validation sets, respectively (**Figure 4A**). Such significantly lower C-indices for EOPE compared to PE are expected, as EOPE cases are usually difficult to predict prognosis. Still, the time-to-delivery prediction for EOPE is on par or better than the prediction of PE diagnosis using the same set of EHR data^33^, demonstrating its potential clinical utility. The KM curves of different predicted survival groups have significant distinctions in both the testing and validation datasets (**Figure 4B** and **4E).** This baseline model consists of the six most important features: gestational age at diagnosis, sPE, PE in a past pregnancy, parity, pulmonary circulatory disorders, and coagulopathies (**Figure 4D**; **Table 3**). All survival plots, dichotomized using the median stratification on each of the 6 features, have log-rank p-values smaller than 0.05 in the discovery dataset (**Supplementary Figure 7)**. Additionally, the AUROCs of binarized classification on delivery in the next 2/7/14 days range from 0.64-0.82 on the testing set (**Figure 4C**) and 0.52-0.68 on the validation set **(Figure 4F).**

**Figure 4:**
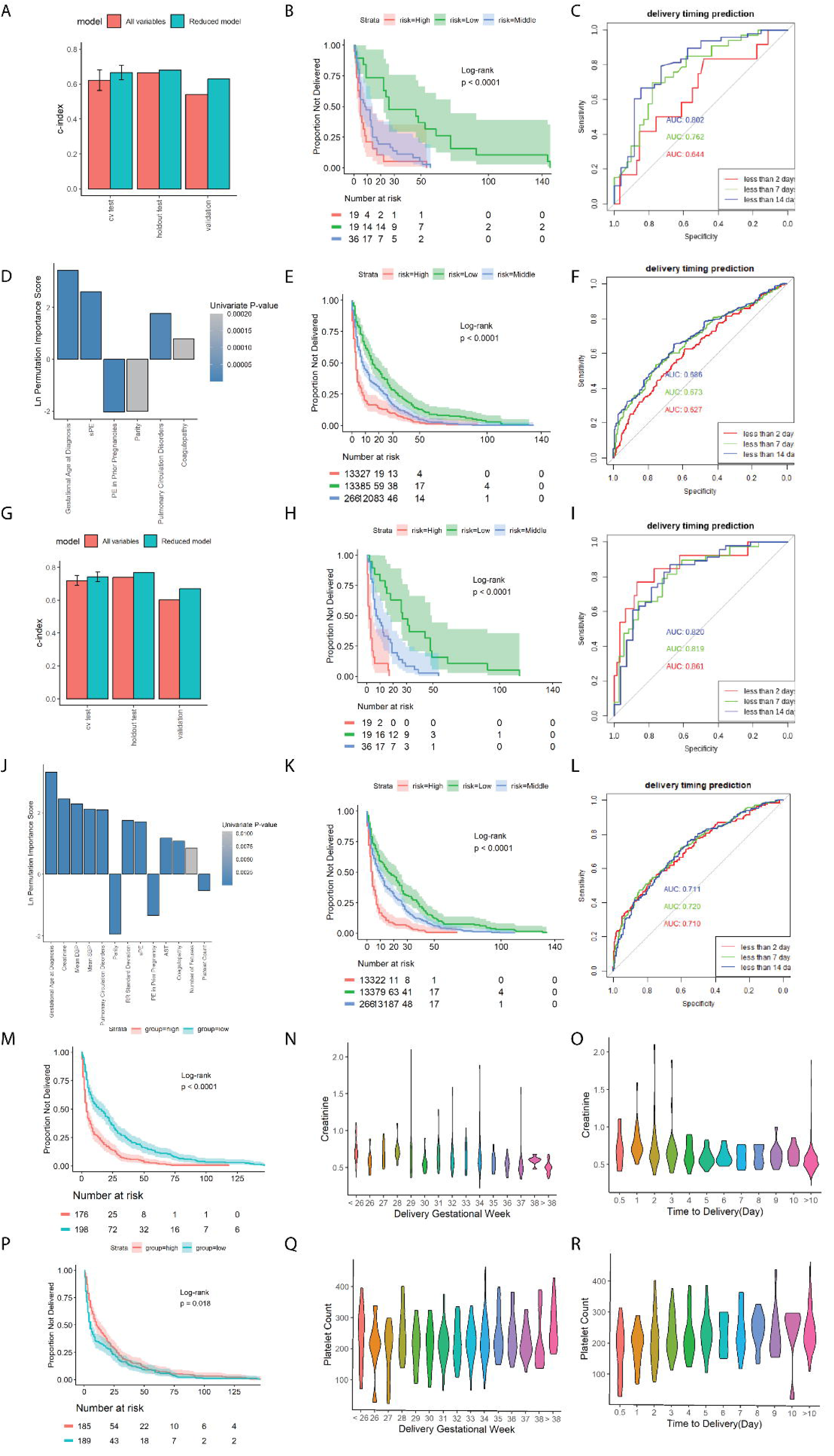
Results, interpretation and evaluation of baseline and full models on the EOPE patient subset. A: The bar plots of C-indices from the original Cox-nnet EOPE baseline model (red) and feature-reduced clinically informative model (green) on the cross-validation and testing set and validation set. B-C: The survival curves of high-risk (top 25%), intermediate-risk (middle 50%) and low-risk groups (bottom 25%), categorized by predicted PI from the reduced EOPE baseline model in A. B, E: hold-out testing data, E: validation data. C, F: ROC curves of prediction delivery time within 2 days, 7 days and 14 days using results from reduced EOPE baseline model (A) on C: hold-out testing data and F: validation data. D: The ln-transformed permutation importance score of features in the EOPE full model. G: The bar plots of C-indices from the original Cox-nnet EOPE full model (red) and its feature-reduced clinically informative model (green) on the cross-validation and testing set and validation set. H, K: The survival curves of high-risk (top 25%), intermediate-risk (middle 50%) and low-risk groups (bottom 25%), categorized by predicted PI from the reduced full model in E. H: hold-out testing data, K: validation data. I, L: ROC curves of prediction delivery time within 2 days, 7 days and 14 days using results from reduced EOPE full model (G) on I: hold-out testing data and L: validation data. J: The ln-transformed permutation importance scores of features in the EOPE full model. M-O: Analysis of creatinine values among the EOPE patients in the discovery cohort. M: The dichotomized survival curves by the median value of creatinine. N, O: Distributions of creatinine values by delivery gestational week (N) and time to delivery (O). P-R: Analysis of platelet counts among the EOPE patients in the discovery cohort. P: The dichotomized survival curves by the median value of platelet counts. Q-R: Distributions of creatinine values by delivery gestational week (Q) and time to delivery (R).

The clinically informative EOPE full model reached much higher accuracy compared to the EOPE baseline model, with median C-indices of 0.74, 0.76, and 0.67 on the cross-validation, testing, and validation sets (**Figure 4G**). The large increases in C-indices are the results of including additional lab tests and blood pressure measurements right around the time of diagnosis of EOPE, confirming their significant clinical values. The 3 risk-stratified groups within the EOPE patient’s cohort also showed significant (log-rank p-value<0.001) differences in the hold-out testing set and validation set (**Figure 4H, 4K).** The AUROCs of chance of delivery in the next 2/7/14 days are significantly improved, ranging from 0.82-0.86 on the testing set (**Figure 4I**) and 0.71-0.72 on the validation set **(Figure 4L).** This model contains 13 important features selected from the original 60 features (**Figure 4J**; **Table 3**). Gestational age at diagnosis continued to be the most important feature. Several other features (eg. PE with severe symptoms, PE in a past pregnancy, parity, and coagulopathy) were of significant importance as well, similar to the EOPE baseline model. Many additional features in the vital signs and lab test categories were also significant, including creatinine value, mean DBP and mean SBP, standard deviation of RR, AST, and platelet counts. Among these 13 features, parity, PE in a prior pregnancy, and higher platelet counts were protective against early delivery **(Figure 4J**).

We created dichotomized survival curves based on creatinine value and platelet count, two new features relative to the EOPE baseline model. Both show strong distinctions between the risk groups (**Figure 4I, 4L**), similar to all other selected features (**Supplementary Figure 7-8**). These two features also revealed systematic trends in associations with the gestational age at delivery and time from diagnosis to delivery. Patients with high creatinine levels were more likely to be delivered within 3 days or less of diagnosis and more likely to deliver preterm (**Figure 4M-4O**). Lower platelet counts were also associated with shorter time to delivery (**Figure 4Q**), even though the platelet levels were not strongly associated with gestational age at delivery among all EOPE patients (**Figure 4R**).

### PE time to deliver predictor graphic user interface (GUI)

To disseminate our model publicly, we packaged the pre-trained clinically informative models above into an interactive, user-friendly web application using R shiny^23^. We named this app “PE time to delivery predictor”. The app contains two main panels: the single-patient prediction panel and the group prediction panel (**Supplementary Figure 9**). The single-patient prediction panel calculates the prognosis index (PI) of a single patient if provided the required clinical variables. The PI score describes the patient’s risk of delivery at the time of the diagnosis of PE, relative to the population. The panel also provides the percentile of the PI score among the training data and displays the results in a histogram figure and a table. The group prediction panel calculates the PI and PI percentile of multiple patients simultaneously and also displays them in a table, below the histogram built on the training data. The app is available at http://garmiregroup.org/PE-prognosis-predictor/app

### Comparison with previous maternal risk prediction models

Lastly, the previously established maternal risk prediction models (i.e. fullPIERS) cannot effectively predict time-to-delivery at the initial diagnosis of PE directly. We calculated the maternal risk of adverse outcomes using the fullPIERS formula on the UM EHR data. We then used this risk score to estimate the time-to-delivery and compared its performance with our proposed model (see **Methods**). The cross-validation C-index of fullPIERS is 0.50±0.005 on all PE patients and 0.60±0.01 on the EOPE subset (**Supplementary Figure 10A**), much worse than those from our models. So are the survival curves grouped by predicted risk (**Supplementary Figure 10B-C**). Thus the time-to-delivery models are not only different but also irreplaceable by the maternal risk prediction models.

## Discussion

PE is a highly heterogenous pregnancy syndrome currently without cure except for delivering the baby and placenta^3,34^. Here we report a new type of survival model to precisely predict the time to delivery as early as the initial diagnosis of PE, subsequent to our recent success in predicting the onset of PE using the same set of EHR data^33^. It helps to save the effort of close monitoring and extensive testing which is conventionally done in resource-rich settings. The simple yet precise models can also be utilized in healthcare systems in resource-limited countries and regions. With such information, clinicians may allocate limited resources in busy antepartum and neonatal ICU beds, or make decisions about the urgency to transfer a patient to a higher level of care in the lack of sufficient resources. As many pregnant women are willing to accept personal risks to improve perceived fetal outcomes, a more concrete model such as the one proposed here will allow them to understand the likely latency and may help them to prepare for delivery emotionally. Many previous studies, such as the fullPIERS and PREP-S models recommended by NICE guidelines, did not predict the precise time of delivery, instead, they fall into very different classification models ^16,18,27,28^ that aim to predict risks of maternal adverse outcomes (**Supplementary Table 5**). Assisting in deciding delivery timing is not their primary purpose. If they were to be used to predict the time-to-delivery directly, the result would not be satisfactory (**Supplementary Figure 10**). Additionally, the prediction window of proposed models is longer than 48 hours in the fullPIERS model, making them good initial assessment tools.

The proposed models confirmed key factors already highlighted in current PE management, including gestational age at the time of diagnosis, sPE, and the use of creatinine, platelet counts and AST as risk factors in clinical guidelines (**Figure 5**)^12–14,18^. This is not surprising, as less time to delivery is likely associated with patients at higher risk for complications based on clinical assessments. However, this class of models also assigns weights of relative importance, among these key factors, a capacity nonexistent in the current ACOG guidelines^12^. Another novel finding is the identification of parity and PE in prior pregnancies as important predictors for delivery timing in all models tested but not included in current guidelines for PE delivery timing (**Figure 5)**. Most importantly, the models predict the timing of delivery at the initial diagnosis and require no more than readily available information from blood work, medical history, and demographics that are routinely collected in medical centers in the US.

**Figure 5:**
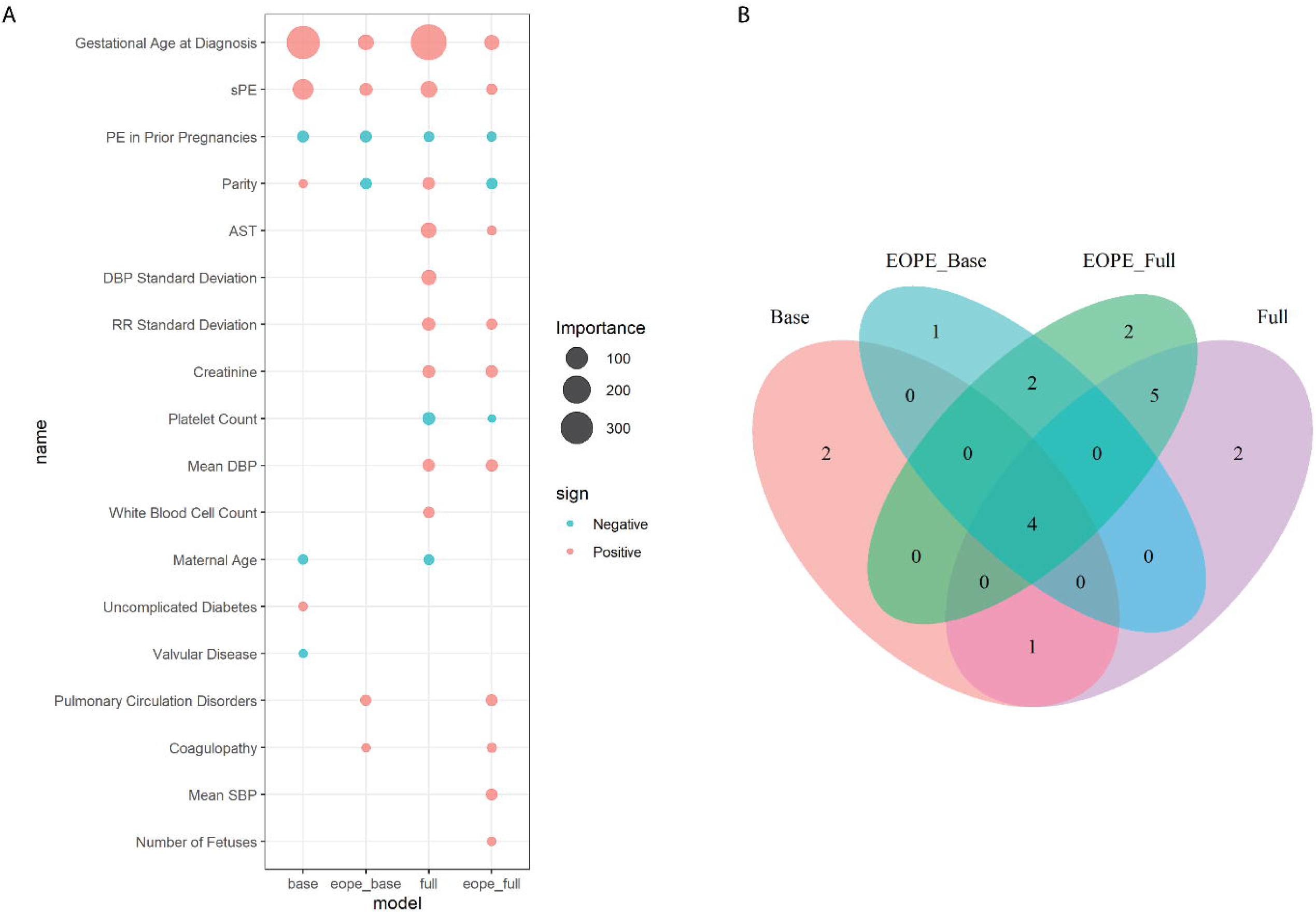
Comparison of important features among the four feature-reduced clinically informative models. A: The bubble plot of important features from PE baseline, EOPE baseline, PE full, and EOPE full models using reduced top important features. The size of the bubbles represents the permutation importance score of each feature. Color represents the sign of features in the time to delivery prediction: a positive sign indicates that a higher value in the feature is associated with a shorter time to delivery and a negative sign means an extension of time to delivery. B: Venn diagram of the important features from the four models shown in A.

There are several noticeable strengths of this study. The models show consistently high performance in survival prediction and classification tasks, better than previous time-to-delivery prediction models using clinical data or biomarkers^37–39^. Unlike the majority of previous studies that are not validated with external data^35,40,41^, our models are validated with an external and independent EHR dataset from UF Health System, despite the noticeable differences between the populations in the two cohorts (**Table 1, 2**). These models also address clinical interpretability by providing importance scores with directionality for each included predictor. Furthermore, the model is designed for accessibility by utilizing fewer than 15 common demographic and disease histories and routinely collected clinical variables in a short observation window. Our approach is much more convenient, as compared to previous studies relying extensively on nonstandard biomarkers such as uterine artery pulsatility index (UtA-PI) or placental growth factor (PLGF)^39–41^. Measurement of these biomarkers is rare in routine prenatal checkups, particularly in lower-income regions, limiting the wide adoption of these biomarker-based models. To maximize the dissemination of the models among clinicians and patients, we have packaged the pre-trained models into a user-friendly shiny application. We aim to embed these models into the EHR system, though it will require additional higher levels of cooperation within the UM Health System. Once integrated, the models will provide clinicians with a fast and accurate assessment of the urgency for delivery at the initial diagnosis of PE.

**Table 2:**
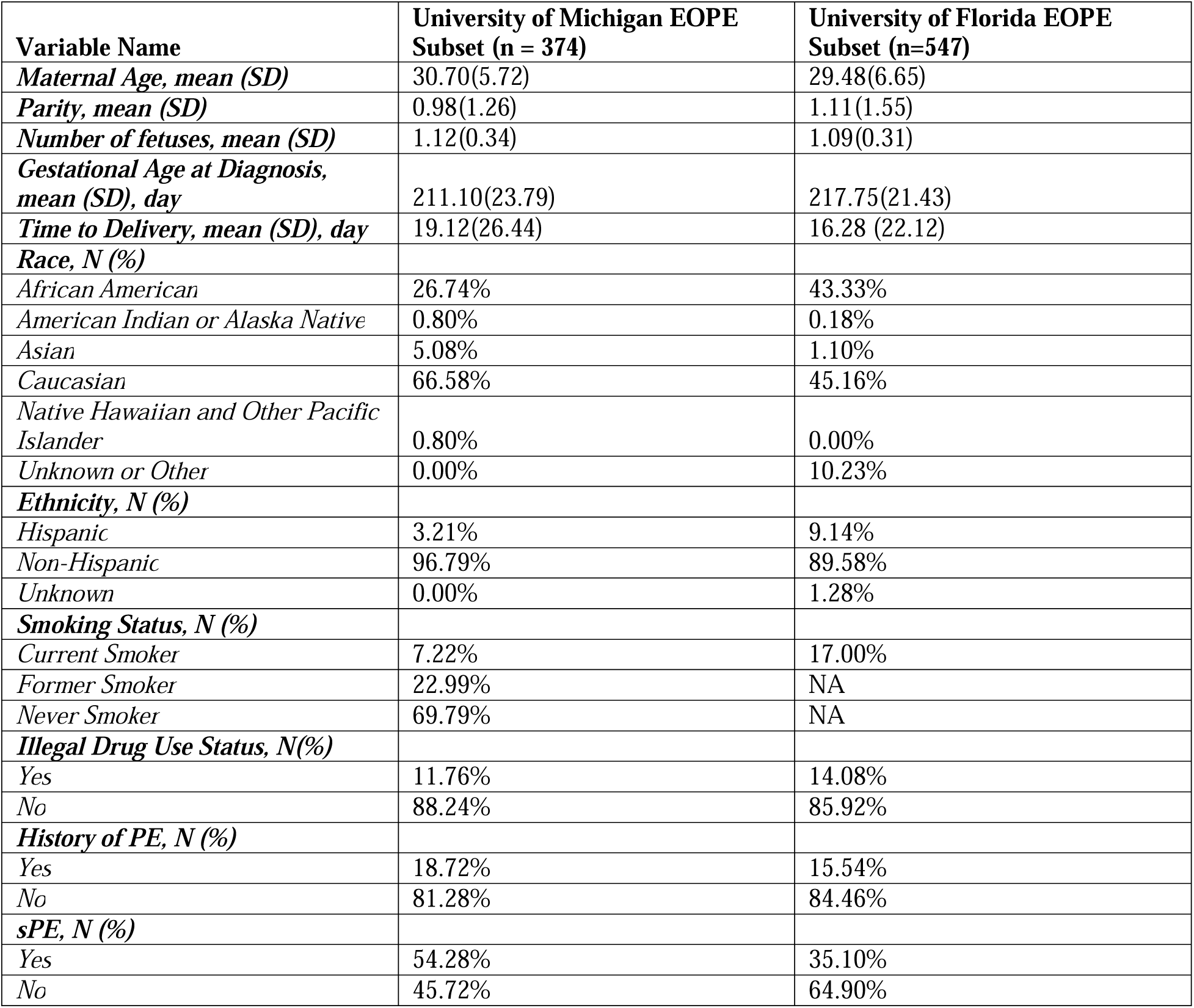
Basic Characteristics of EOPE patients in this study.

| **Variable Name** | **University of Michigan EOPE**<br>**Subset (n = 374)** | **University of Florida EOPE**<br>**Subset (n=547)** |
| --- | --- | --- |
| *Maternal Age, mean (SD)* | 30.70(5.72) | 29.48(6.65) |
| *Parity, mean (SD)* | 0.98(1.26) | 1.11(1.55) |
| *Number of fetuses, mean (SD)* | 1.12(0.34) | 1.09(0.31) |
| *Gestational Age at Diagnosis,*<br>*mean (SD), day* | 211.10(23.79) | 217.75(21.43) |
| *Time to Delivery, mean (SD), day* | 19.12(26.44) | 16.28 (22.12) |
| **Race, N (%)** |  |  |
| *African American* | 26.74% | 43.33% |
| *American Indian or Alaska Native* | 0.80% | 0.18% |
| *Asian* | 5.08% | 1.10% |
| *Caucasian* | 66.58% | 45.16% |
| *Native Hawaiian and Other Pacific*<br>*Islander* | 0.80% | 0.00% |
| *Unknown or Other* | 0.00% | 10.23% |
| **Ethnicity, N (%)** |  |  |
| *Hispanic* | 3.21% | 9.14% |
| *Non-Hispanic* | 96.79% | 89.58% |
| *Unknown* | 0.00% | 1.28% |
| **Smoking Status, N (%)** |  |  |
| *Current Smoker* | 7.22% | 17.00% |
| *Former Smoker* | 22.99% | NA |
| *Never Smoker* | 69.79% | NA |
| **Illegal Drug Use Status, N(%)** |  |  |
| *Yes* | 11.76% | 14.08% |
| *No* | 88.24% | 85.92% |
| **History of PE, N (%)** |  |  |
| *Yes* | 18.72% | 15.54% |
| *No* | 81.28% | 84.46% |
| **sPE, N (%)** |  |  |
| *Yes* | 54.28% | 35.10% |
| *No* | 45.72% | 64.90% |

**Table 3:**
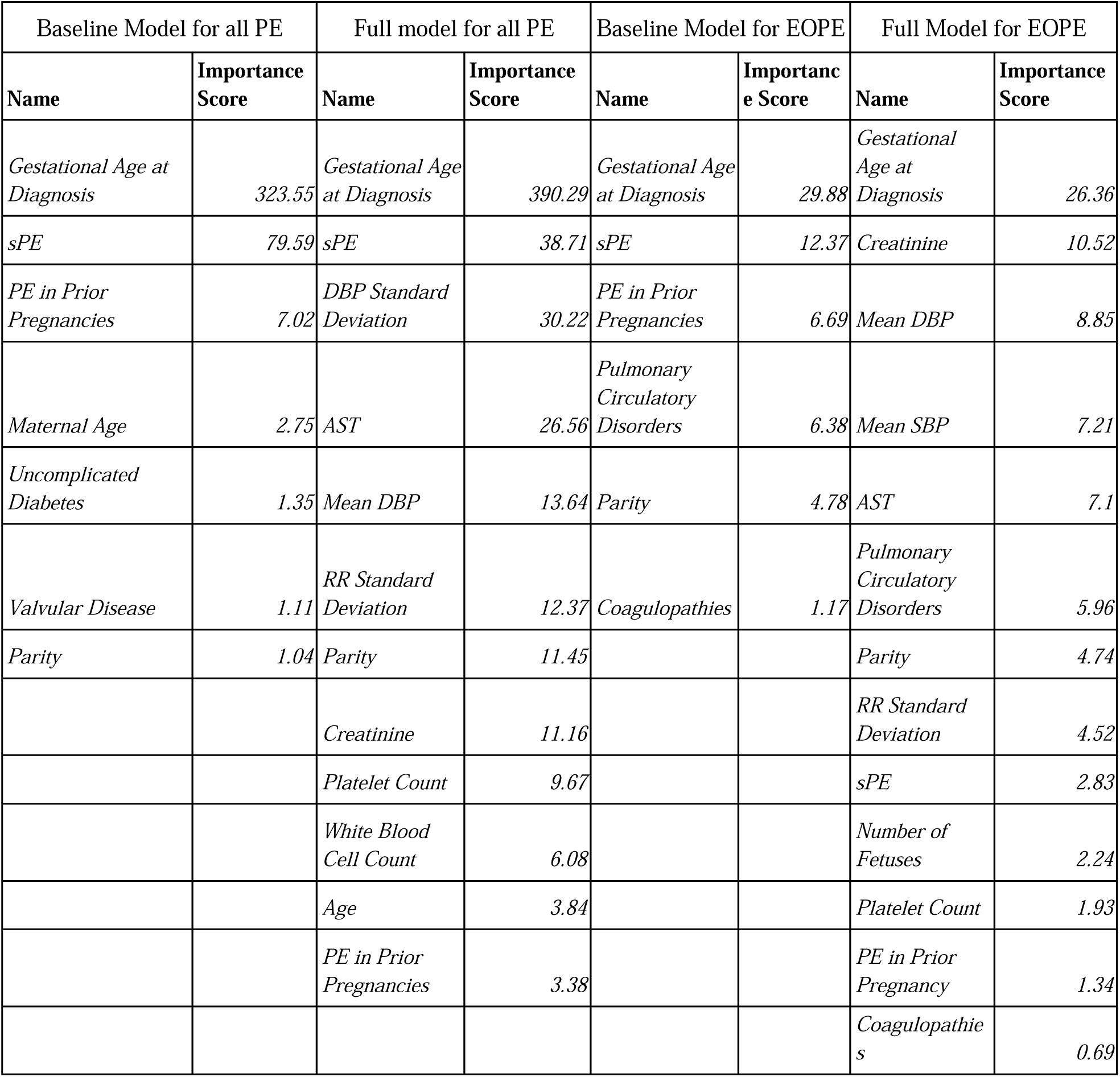
Features and their permutation importance score in each reduced model.

A few caveats to this study are potentially limiting. Firstly, the ICD coding system lags behind the most recent diagnostic guidelines of PE. However, the codes that are entered into the EHR are based on the clinical assessments of the treating physicians at two academic medical centers, therefore they most likely reflect contemporary diagnostic standards. Also as a retrospective study, the delivery timing can be influenced by clinicians’ previous judgment, changes in hospital protocol, communication between patient and provider, intensive care resource availability, and each patient’s intentions. However, since these models generally perform well on the external validation set as they achieve C-indices of 0.7 or even close to 0.80, we believe that these subjective factors may not be the main concerns for achieving high prediction power, rather, additional data modalities may help. Prospective investigations of this model’s performance in other medical centers would be necessary to confirm the findings. Lastly, our data came from two medical centers with high levels of obstetrics care and therefore testing the model in other settings (eg. other countries and rural regions) will be deemed valuable.

In summary, we have developed the first accurate, deep-learning-based, time-to-delivery prediction models for PE and EOPE patients. The models are disseminated with an easy-to-use web app. Adoption of these models could provide clinicians and patients with valuable management plans to predict and prepare for the best delivery time of each PE pregnancy. Further prospective investigation of the performance of these models is necessary to provide feedback and potential improvement of these models.

## Acknowledgment

We thank Anisa Driscoll and Kate Smith from the University of Michigan Precision Health for providing technical support when extracting data used in this study. We acknowledge the University of Florida Integrated Data Repository (IDR) and the UF Health Office of the Chief Data Officer for providing the analytic data set for this project.

## Data Sharing Statement

We are unable to publicly share electronic health records data due to its potential to reveal sensitive patient information. However, interested investigators who met the criteria for accessing sensitive data can contact the University of Michigan Precision Health’s Research Scientific Facilitators at (also see https://research.medicine.umich.edu/our-units/data-office-clinical-translational-research/data-access) to inquire about the UM dataset and the necessary steps regarding ethics committee approval and data sharing agreement. Please contact the UF Health Integrated Data Repository (IDR, https://idr.ufhealth.org/) at to inquire about the UF dataset and the necessary steps regarding ethics committee approval and data sharing agreement.

## Code Availability

Codes used for analysis are available at https://github.com/lanagarmire/PE_delivery

## Author’s Contribution

LG conceived this project and supervised the study, after discussing it with ESL. XY conducted data analysis, implemented the Shiny app, and wrote the manuscript. HKB, ADM, KX, and DJL collaborated on validation using the UF cohort. ESL and ADM provided clinical assessments and assistance. DG assisted with Shiny app editing and troubleshooting. All authors have read, revised, and approved the manuscript.

## Abbreviations

PE: preeclampsia
sPE: Preeclampsia with severe features
EOPE: early-onset preeclampsia
LOPE: late-onset preeclampsia
EHR: electronic health record
SBP: systolic blood pressure
DBP: diastolic blood pressure
RR: respiratory rate
HELLP: hemolysis, elevated liver enzymes, low platelet count
AST: aspartate transaminase
PI: prognosis score
UM: University of Michigan
UF: University of Florida
ICD-10: The International Classification of Diseases, Tenth Revision
MAP: mean arterial pressure
UtA-PI: uterine artery pulsatility index
PLGF: placental growth factor
ACOG: American College of Obstetricians and Gynecologist
AUROC: Area Under the Receiver-Operator curve

