## Supplementary materials for "Deep learning-based prognosis models accurately predict the time to delivery among preeclampsia patients using health records at the time of diagnosis"

**Supplementary Table**

**Supplementary Table 1: ICD diagnosis codes used to select PE cohort**

| ***ICD diagnosis code*** | ***Diagnosis Description*** |
| --- | --- |
| *O14.03* | *Mild to moderate pre-eclampsia, third trimester* |
| *O14.02* | *Mild to moderate pre-eclampsia, second trimester* |
| *O14.93* | *Unspecified pre-eclampsia, third trimester* |
| *O14.92* | *Unspecified pre-eclampsia, second trimester* |
| *O14.13* | *Severe pre-eclampsia, third trimester* |
| *O14.12* | *Severe pre-eclampsia, second trimester* |
| *642.43* | *Mild or unspecified pre-eclampsia, antepartum condition or complication* |
| *642.53* | *Severe pre-eclampsia, antepartum condition or complication* |

**Supplementary Table 2**: The list of all (60) features, including baseline features and additional features used in this study

| ***Baseline Features*** | | ***Additional Features*** |
| --- | --- | --- |
| ***PE information at diagnosis*** | ***Comorbidities*** | ***Lab Results*** |
| *Number of fetuses* | *Alcohol Abuse* | *Creatinine* |
| *Diagnosis Gestational age (days)* | *Blood Loss Anemia* | *Mean_Corpuscular_Hgb* |
| *Time to deliver(days)* | *Cardiac Arrhythmias* | *Mean_Corpuscular_Hgb_Conc* |
| *Severe PE(sPE)* | *Chronic Pulmonary Disease* | *Hematocrit* |
|  | *Coagulopathy* | *Red_Cell_Distribution_Width* |
| ***Demographics*** | *Congestive Heart Failure* | *White_Blood_Cell_Count* |
| *Current Smoker* | *Deficiency Anemia* | *Mean_Platelet_Volume* |
| *Former Smoker* | *Depression* | *Platelet_Count* |
| *Illegal Drug User* | *Diabetes Complicated* | *AST* |
| *African American* | *Diabetes Uncomplicated* |  |
| *Asian* | *Drug Abuse* | ***Vital Signs*** |
| *Hispanic* | *Fluid Electrolyte Disorders* | *Mean Diastolic BP* |
| *Age* | *Hypothyroidism* | *Diastolic BP Standard Deviation* |
|  | *Liver Disease* | *Mean Systolic BP* |
| ***Pregnancy related medical history*** | *Metastatic Cancer* | *Systolic BP Standard Deviation* |
| *Parity* | *Obesity* | *Mean Respiratory Rate (RR)* |
| *PE in prior pregnancy* | *Other Neurological Disorders* | *RR Standard Deviation* |
| *C-section in prior pregnancy* | *Peptic Ulcer Disease Excluding Bleeding* |  |
| *History of Renal disease* | *Peripheral VascularDisorders* |  |
| *History of Gestational Diabetes* | *Psychoses* |  |
| *History of Abruption* | *Pulmonary Circulation Disorders* |  |
| *Preterm Labor in prior pregnancy* | *Renal Failure* |  |
| ***Fetal Development*** | *Rheumatoid Arthritis Collagen Vascular Diseases* |  |
| *poor fetal development* | *Solid Tumor Without Metastasis* |  |
|  | *Valvular Disease* |  |
|  | *Weight Loss* |  |

**Supplementary Table 3: Analysis of Deviance of each feature included in the final models represented by Chi-square**

| baseline | | | full | | | EOPE base | | | EOPE full | | |
| --- | --- | --- | --- | --- | --- | --- | --- | --- | --- | --- | --- |
| name | chisq | pval | name | chisq | pval | name | chisq | pval | name | chisq | pval |
| diag_GA | 689.95 | 4.58E-152 | diag_GA | 689.95 | 4.58E-152 | diag_GA | 72.43 | 1.73E-17 | BPDiaMean | 86.07 | 1.74E-20 |
| past_pe | 48.3 | 3.65E-12 | BPDiaSD | 112.75 | 2.45E-26 | SeverePE | 31.88 | 1.64E-08 | BPSysMean | 84.44 | 3.97E-20 |
| SeverePE | 44.5 | 2.55E-11 | RRSD | 72.26 | 1.88E-17 | past_pe | 27.2 | 1.84E-07 | diag_GA | 72.43 | 1.73E-17 |
| Parity | 27.65 | 1.45E-07 | creatinine_value | 52.68 | 3.92E-13 | Parity | 15.67 | 7.53E-05 | creatinine_value | 50.66 | 1.10E-12 |
| DiabetesUncomplicated | 6.32 | 0.011935 | AST | 52 | 5.54E-13 | PulmonaryCirculationDisorders | 12.69 | 0.000368 | SeverePE | 31.88 | 1.64E-08 |
| ValvularDisease | 5.08 | 0.024157 | past_pe | 48.3 | 3.65E-12 | Coagulopathy | 12.15 | 0.000492 | past_pe | 27.2 | 1.84E-07 |
| age | 4.54 | 0.033195 | SeverePE | 44.5 | 2.55E-11 |  |  |  | AST | 24.65 | 6.89E-07 |
|  |  |  | BPDiaMean | 42.52 | 7.00E-11 |  |  |  | Platelet_Count | 16.94 | 3.87E-05 |
|  |  |  | Platelet_Count | 36.29 | 1.70E-09 |  |  |  | Parity | 15.67 | 7.53E-05 |
|  |  |  | Parity | 27.65 | 1.45E-07 |  |  |  | RRSD | 14.99 | 0.000108 |
|  |  |  | White_Blood_Cell_Count | 9.5 | 0.002054 |  |  |  | Pulmonary Circulation Disorders | 12.69 | 0.000368 |
|  |  |  | age | 4.54 | 0.033195 |  |  |  | Coagulopathy | 12.15 | 0.000492 |
|  |  |  |  |  |  |  |  |  | NUMBER_OF_FETUSES | 5.82 | 0.015805 |

**Supplementary Table 4: TRIPOD checklist: Prediction model development and validation**

| **Section/Topic** | **Item** |  | **Checklist Item** | **Page** |
| --- | --- | --- | --- | --- |
| **Title and abstract** | | | | |
| Title | 1 | D;V | Identify the study as developing and/or validating a multivariable prediction model, the target population, and the outcome to be predicted. | 1 |
| Abstract | 2 | D;V | Provide a summary of objectives, study design, setting, participants, sample size, predictors, outcome, statistical analysis, results, and conclusions. | 3 |
| **Introduction** | | | | |
| Background and objectives | 3a | D;V | Explain the medical context (including whether diagnostic or prognostic) and rationale for developing or validating the multivariable prediction model, including references to existing models. | 6 |
|  | 3b | D;V | Specify the objectives, including whether the study describes the development or validation of the model or both. | 7 |
| **Methods** | | | | |
| Source of data | 4a | D;V | Describe the study design or source of data (e.g., randomized trial, cohort, or registry data), separately for the development and validation data sets, if applicable. | 7 |
|  | 4b | D;V | Specify the key study dates, including start of accrual; end of accrual; and, if applicable, end of follow-up. | 7 |
| Participants | 5a | D;V | Specify key elements of the study setting (e.g., primary care, secondary care, general population) including number and location of centres. | 8 |
|  | 5b | D;V | Describe eligibility criteria for participants. | 8 |
|  | 5c | D;V | Give details of treatments received, if relevant. | NA |
| Outcome | 6a | D;V | Clearly define the outcome that is predicted by the prediction model, including how and when assessed. | 9 |
|  | 6b | D;V | Report any actions to blind assessment of the outcome to be predicted. | NA |
| Predictors | 7a | D;V | Clearly define all predictors used in developing or validating the multivariable prediction model, including how and when they were measured. | 8; Supp 1 |
|  | 7b | D;V | Report any actions to blind assessment of predictors for the outcome and other predictors. | NA |
| Sample size | 8 | D;V | Explain how the study size was arrived at. | 8 |
| Missing data | 9 | D;V | Describe how missing data were handled (e.g., complete-case analysis, single imputation, multiple imputation) with details of any imputation method. | 8; sup p1 |
| Statistical analysis methods | 10a | D | Describe how predictors were handled in the analyses. | 8 |
|  | 10b | D | Specify type of model, all model-building procedures (including any predictor selection), and method for internal validation. | 8-9 |
|  | 10c | V | For validation, describe how the predictions were calculated. | Sup p2 |
|  | 10d | D;V | Specify all measures used to assess model performance and, if relevant, to compare multiple models. | 16 |
|  | 10e | V | Describe any model updating (e.g., recalibration) arising from the validation, if done. | NA |
| Risk groups | 11 | D;V | Provide details on how risk groups were created, if done. | 9 |
| Development vs. validation | 12 | V | For validation, identify any differences from the development data in setting, eligibility criteria, outcome, and predictors. | Table1 |
| **Results** | | | | |
| Participants | 13a | D;V | Describe the flow of participants through the study, including the number of participants with and without the outcome and, if applicable, a summary of the follow-up time. A diagram may be helpful. | Figure 1 |
|  | 13b | D;V | Describe the characteristics of the participants (basic demographics, clinical features, available predictors), including the number of participants with missing data for predictors and outcome. | Table 1; sup figure 2 |
|  | 13c | V | For validation, show a comparison with the development data of the distribution of important variables (demographics, predictors and outcome). | Table 1; sup figure 1 |
| Model development | 14a | D | Specify the number of participants and outcome events in each analysis. | 10 |
|  | 14b | D | If done, report the unadjusted association between each candidate predictor and outcome. | Sup table 3-6 |
| Model specification | 15a | D | Present the full prediction model to allow predictions for individuals (i.e., all regression coefficients, and model intercept or baseline survival at a given time point). | Figure 2-4; sup table 3-6 |
|  | 15b | D | Explain how to the use the prediction model. | 10, 15, Shiny app |
| Model performance | 16 | D;V | Report performance measures (with CIs) for the prediction model. | Figure 2-4 |
| Model-updating | 17 | V | If done, report the results from any model updating (i.e., model specification, model performance). | NA |
| **Discussion** | | | | |
| Limitations | 18 | D;V | Discuss any limitations of the study (such as nonrepresentative sample, few events per predictor, missing data). | 17 |
| Interpretation | 19a | V | For validation, discuss the results with reference to performance in the development data, and any other validation data. | 11-15 |
|  | 19b | D;V | Give an overall interpretation of the results, considering objectives, limitations, results from similar studies, and other relevant evidence. | 15-17 |
| Implications | 20 | D;V | Discuss the potential clinical use of the model and implications for future research. | 17 |
| **Other information** | | | | |
| Supplementary information | 21 | D;V | Provide information about the availability of supplementary resources, such as study protocol, Web calculator, and data sets. | Sup materials |
| Funding | 22 | D;V | Give the source of funding and the role of the funders for the present study. | 17 |

*Items relevant only to the development of a prediction model are denoted by D, items relating solely to a validation of a prediction model are denoted by V, and items relating to both are denoted D;V. We recommend using the TRIPOD Checklist in conjunction with the TRIPOD Explanation and Elaboration document.

**Supplementary Table 5: Comparison of previous models on PE delivery timing**

| **Name** | **Outcome of interest** | **prediction window** | **model type** |
| --- | --- | --- | --- |
| fullPIERS | risk of maternal adverse outcomes | most accurate in 48 hours;  less accurate in 7 days | logistic regression |
| miniPIERS | risk of maternal adverse outcomes | 48 hours after diagnosis | logistic regression |
| PREP-L | risk of maternal adverse outcomes | diagnosis to discharge | logistic regression |
| PREP-S | time of maternal adverse outcomes | 48 hours, 7 days after diagnosis and by discharge | parametric survival model |
| Schmidt et al. | risk of maternal adverse outcomes | Any visit to 14 days after delivery | tree-based machine learning models |
| Proposed models | delivery time that minimizes maternal and fetal risk from historical decisions | Diagnosis to delivery | deep learning model |

**Supplementary Figures**

**Supplementary Figure 1**: Cox-nnet Architecture

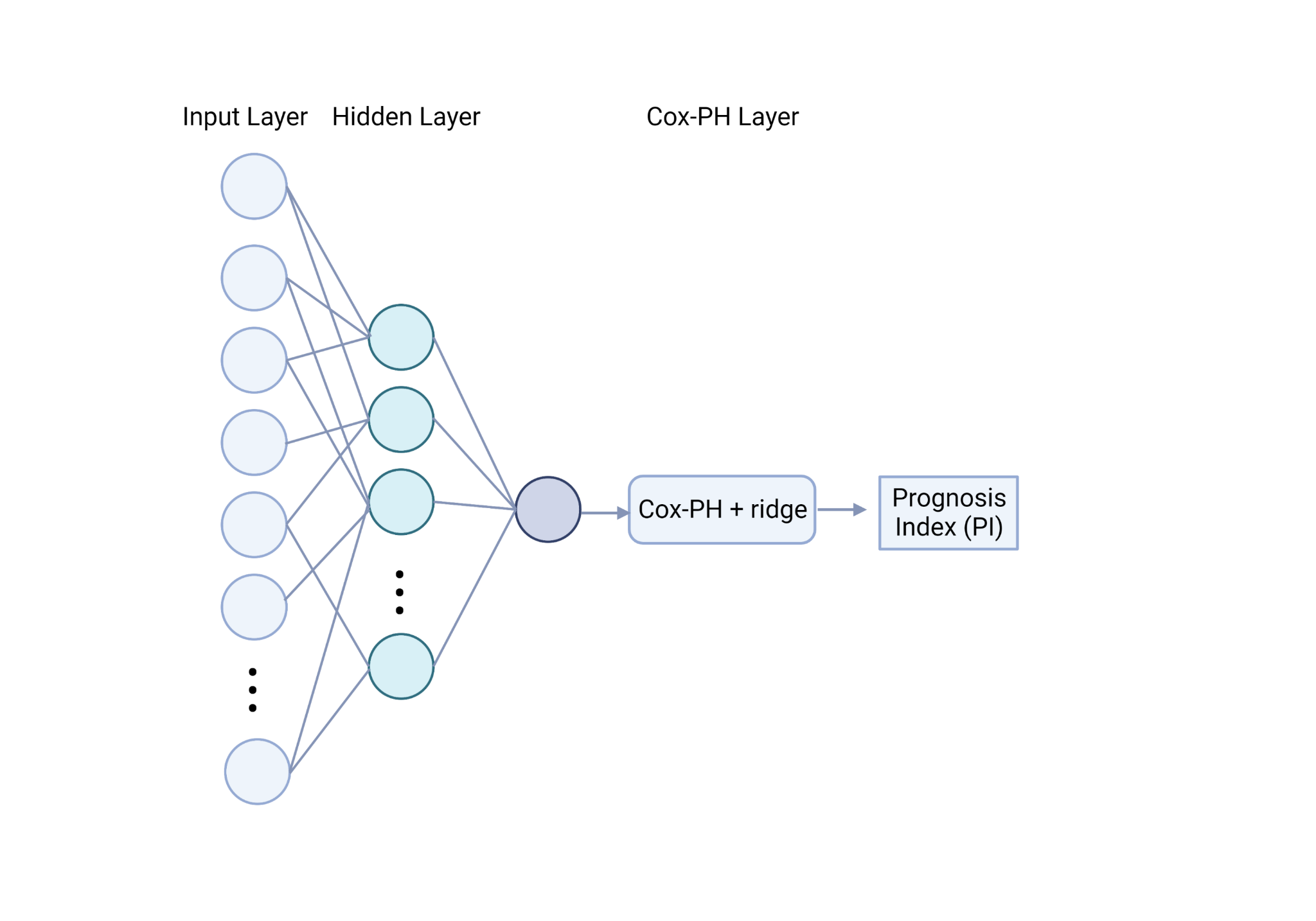

**Supplementary Figure 2:** A) Number of measurement distribution and median time between diagnosis and measurement distribution for each lab variable;

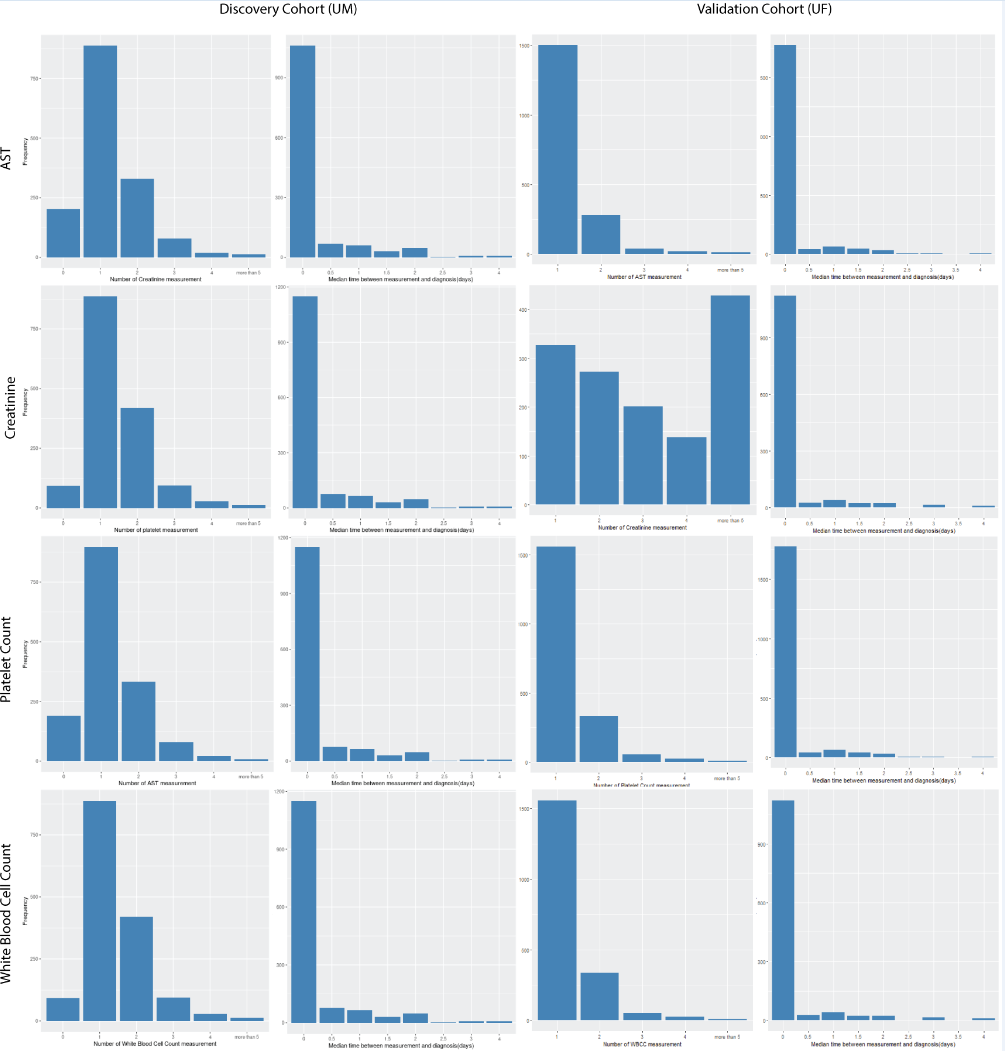

B) Correlation heatmap of all features

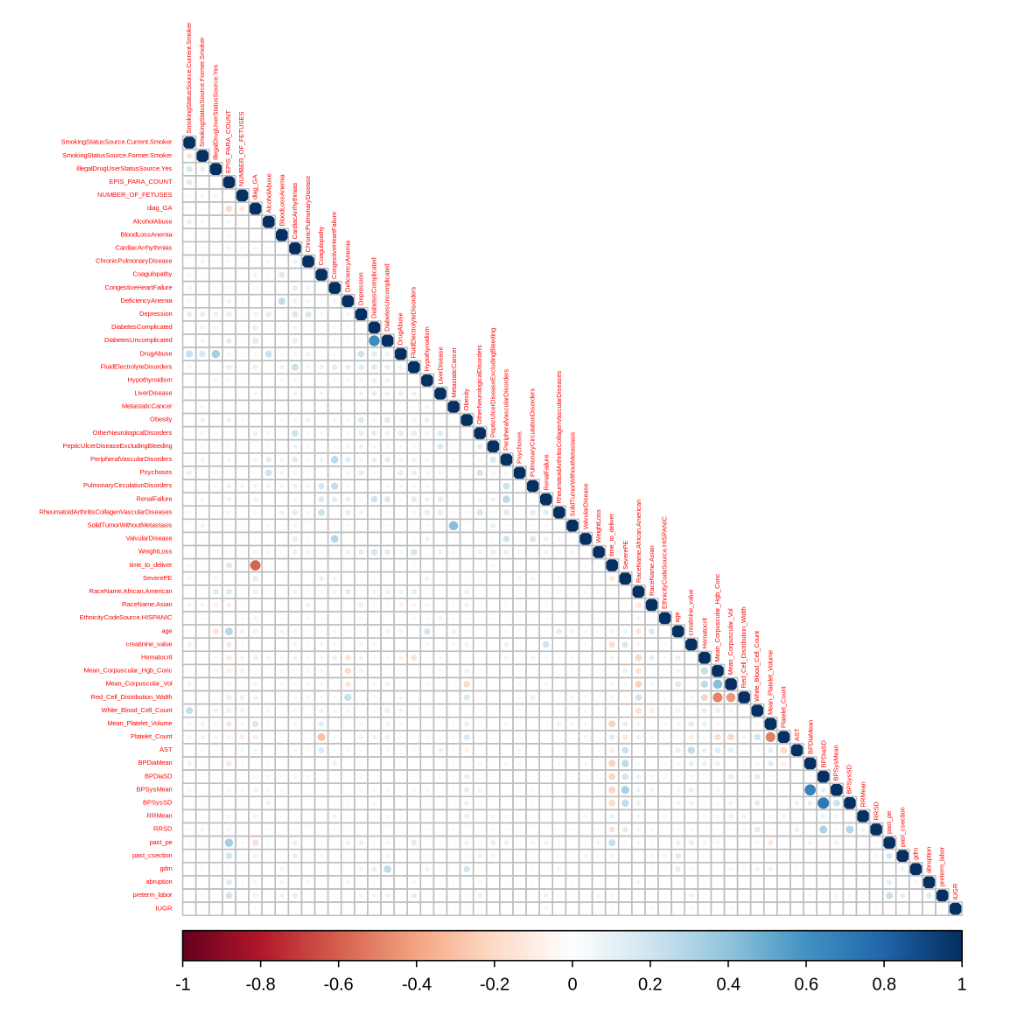

**Supplementary Figure 3:** Dichotomized survival curves by the median value of each important feature in the PE baseline model;
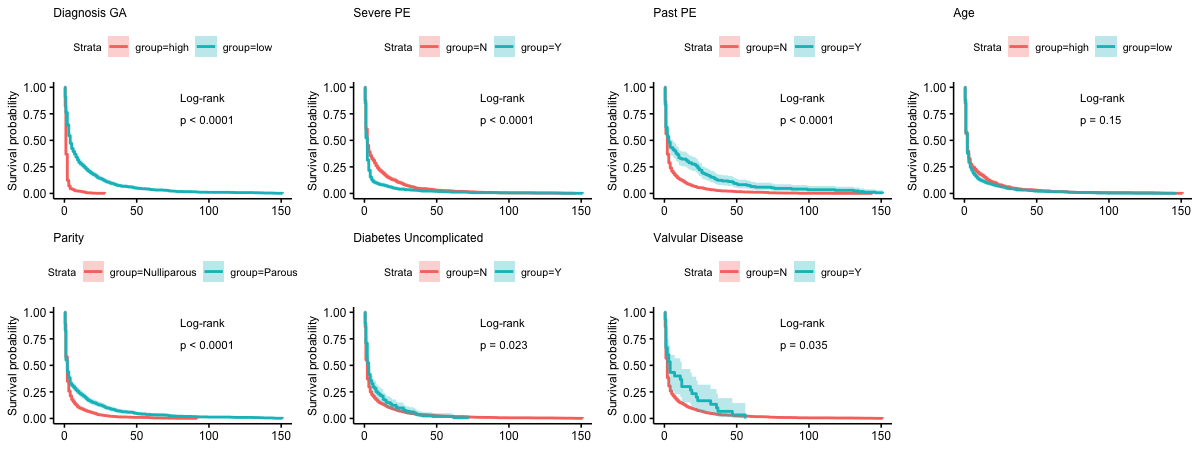

**Supplementary Figure 4: Evaluation of model build on preterm patients (less than 37 weeks of gestation).** Plots A-D show the C-index, Kaplan-Meier curves, AUC scores and feature importance scores of the baseline model and plots E-H are the same metrics of the full model.

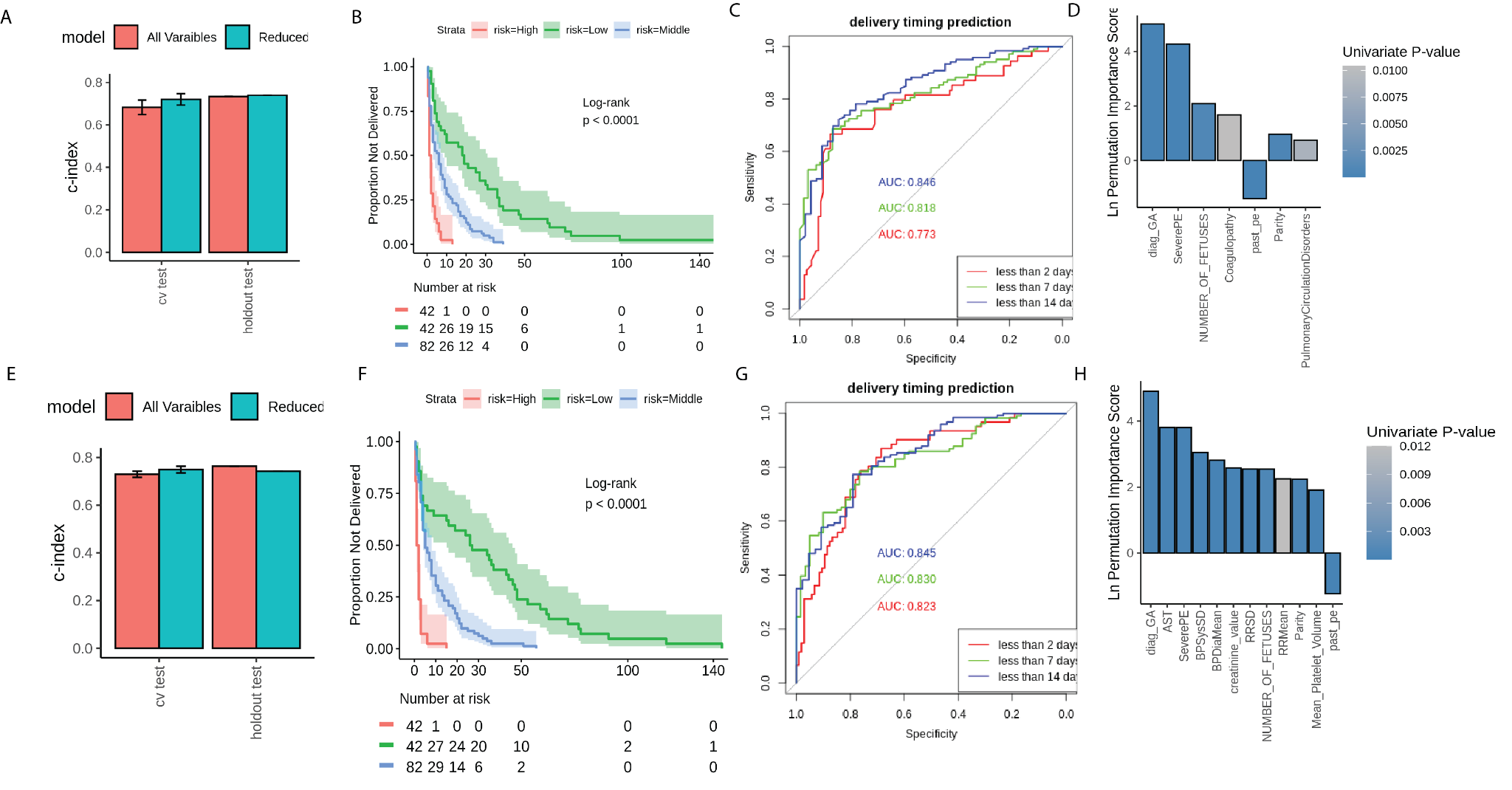

**Supplementary Figure 5**: Dichotomized survival curves by the median value of each important feature in the PE full model

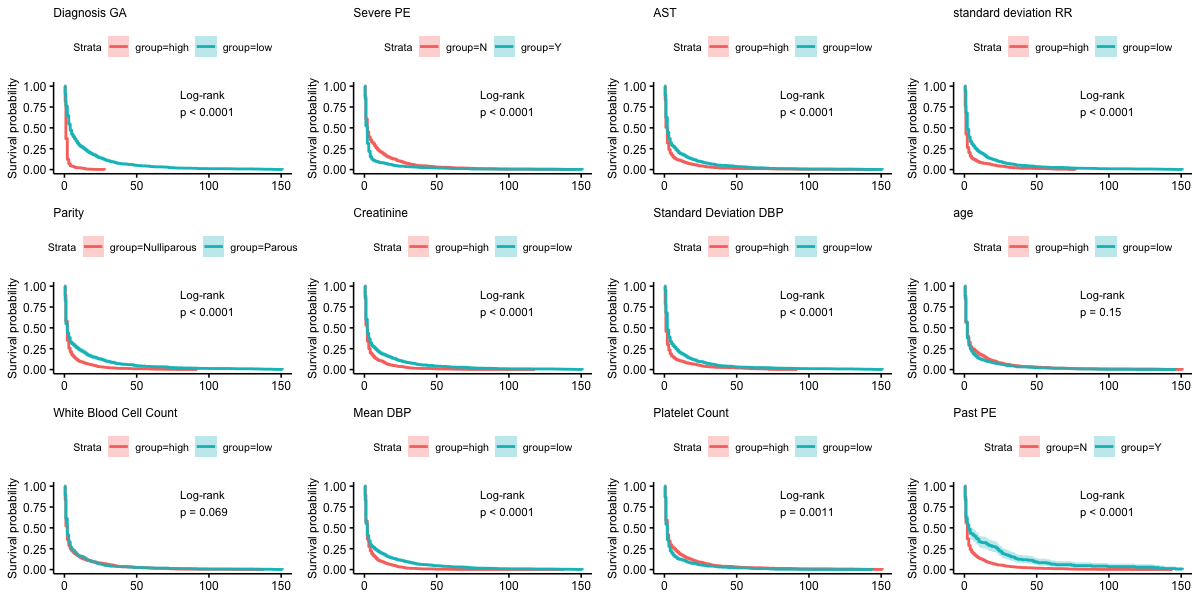

**Supplementary Figure 6**: Distribution of AST, the standard deviation of DBP and the standard deviation of RR over delivery gestational age

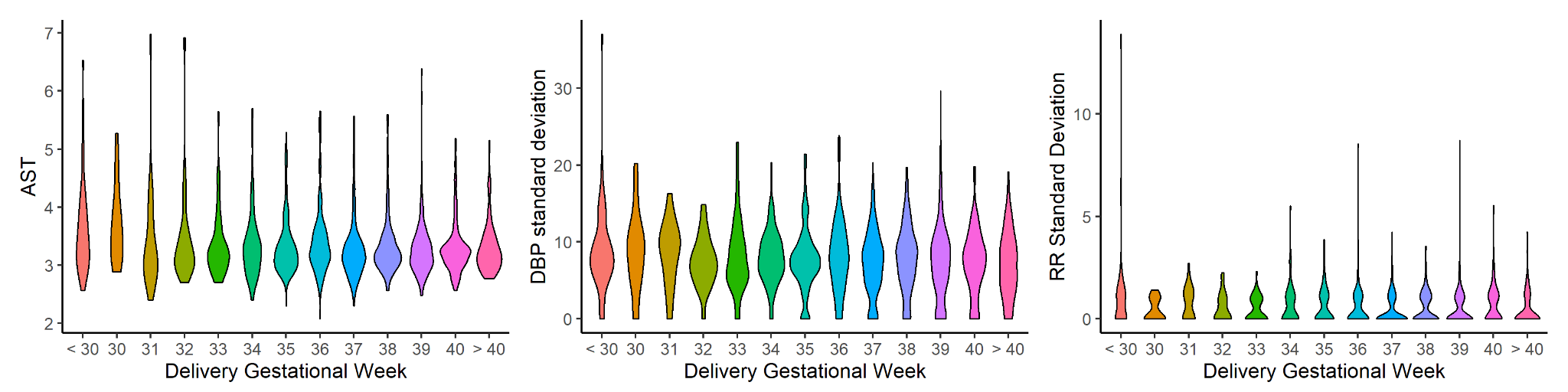

**Supplementary Figure 7:** Dichotomized survival curves by the median value of each important feature in the EOPE baseline model.
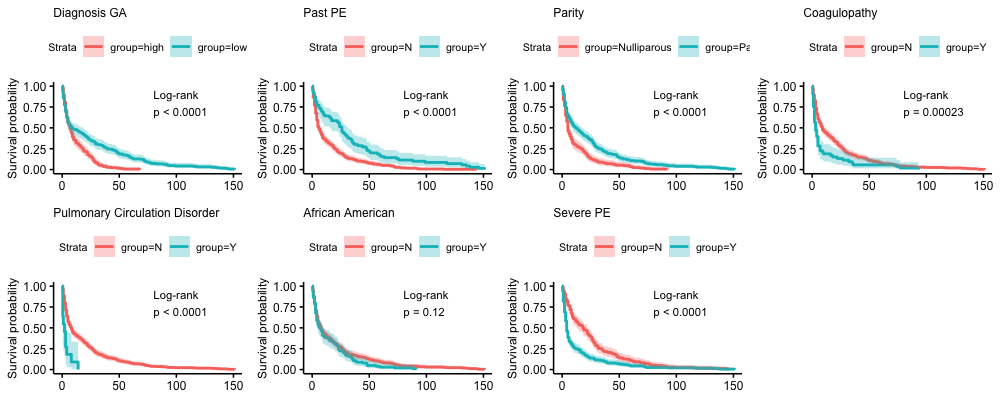

**Supplementary Figure 8:** Dichotomized survival curves by the median value of each important feature in the EOPE full model

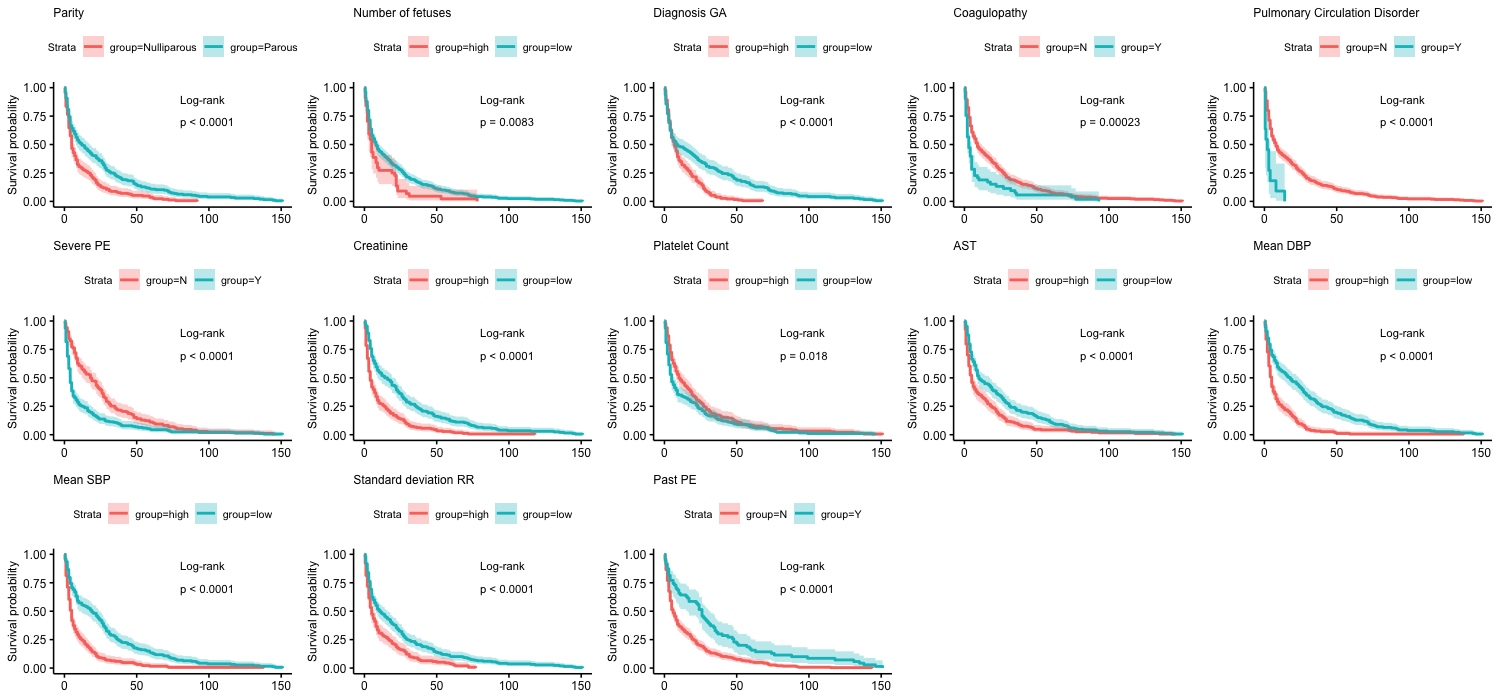

**Supplementary Figure 9:** A snapshot of the Rshiny Interactive Graph User Interface of the PE time-to-delivery predictor.

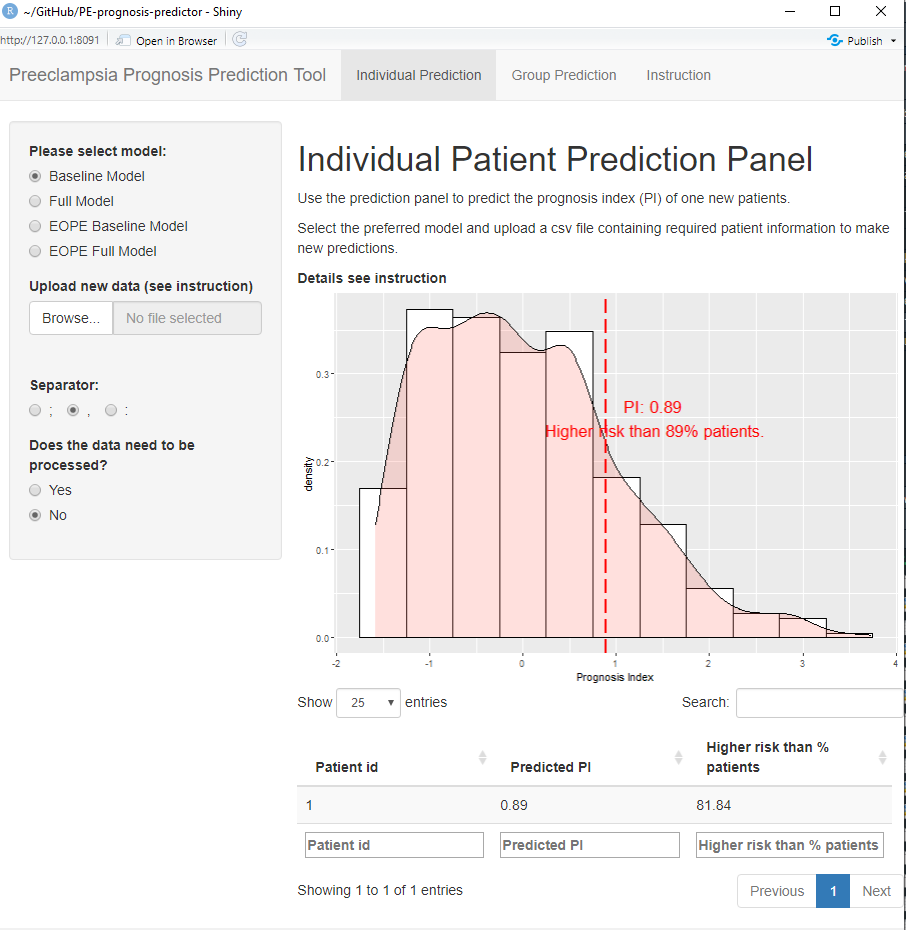

**Supplementary Figure 10: Attempting to predict time to delivery using fullPIERS model.** A) 5 fold CV c-index by applying fullPIERS on all PE patients and EOPE patients. B-C) Survival curves grouped by fullPIERS estimated risk for all PE patients (B) and EOPE patients(C)

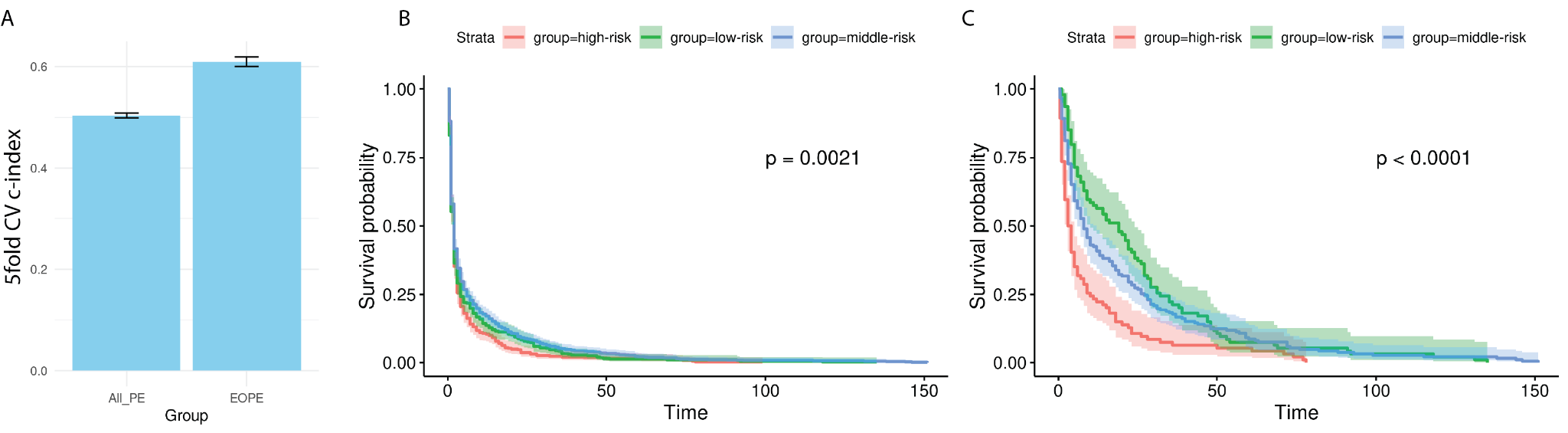
